# Impact of sensitive circulating tumor DNA monitoring on CT scan intervals during postoperative colorectal cancer surveillance

**DOI:** 10.1101/2022.09.03.22279571

**Authors:** Tomoko Sasaki, Takeshi Iwaya, Mizunori Yaegashi, Masashi Idogawa, Hayato Hiraki, Masakazu Abe, Yuka Koizumi, Noriyuki Sasaki, Akiko Yashima-Abo, Ryosuke Fujisawa, Fumitaka Endo, Shoichiro Tange, Tomomi Hirano, Koki Otsuka, Akira Sasaki, Mari Masuda, Masashi Fujita, Hidewaki Nakagawa, Fumiaki Takahashi, Yasushi Sasaki, Takashi Tokino, Satoshi S. Nishizuka

## Abstract

**Objective:** This study investigated whether digital PCR (dPCR)-based circulating tumor DNA (ctDNA) monitoringcan allow longer intervals between computed tomography (CT) scans during postoperative surveillance of colorectal cancer (CRC).

**Design:** The longitudinal dynamics of ctDNA for 52 patients with CRC as measured by dPCR using probes targeting 87 individual tumor-specific mutations (1-5 per patient) were compared with results from conventional (i.e., clinical) surveillance using serum tumor markers and CT. A total of 382 CT procedures were carried out for the patient cohort (3.3/year per patient) and the median lead time from ctDNA relapse to clinical relapse was 182 days (range 0-376 days). If the CT interval was annual, potential delays in detection of clinical relapse would have occurred for 7 of the 10 patients who experienced clinical relapse (9 of 13 events), with a median delay of 164 days (range, 0-267 days). If annual CT surveillance was performed together with ctDNA monitoring, 218 (57.1%) CTs would not have been needed to detect the first clinical relapse. Nonetheless, ctDNA monitoring would still have provided a lead time of 339 days for detection of clinical relapse (range, 42-533 days).

**Conclusion:** Our findings suggest that the ctDNA monitoring as part of post-operative surveillance and clinical relapse detection for patients with CRC could allow the CT interval to be lengthened.

## INTRODUCTION

In 2020, over 1.8 million new cases and 915,000 deaths due to colorectal cancer (CRC) were reported worldwide.^1^ Approximately two-thirds of patients with stage II or III CRC undergo resection with curative intent.^2^ For patients with relapse, resection of metastatic sites followed by systemic chemotherapy have been shown to improve outcomes.^3, 4^ A main goal of postoperative surveillance as part of current therapeutic strategies is to improve disease-specific and overall survival (OS) through early detection of relapse and timely treatment interventions. During the 2000s, results of several meta-analyses suggested that intensive follow-up after CRC resection with curative intent could prolong OS and reduce the re-resection rate for recurrent disease.^5–7^ However, neither data from randomized trials nor a large cohort study conducted in the 2010s showed that intensive surveillance for patients with CRC provided significant benefit.^8–11^ In the 2020s, practical guidelines still recommend intensive surveillance including a computed tomography scan (CT) every 6-12 months and serum carcinoembryonic antigen (CEA) testing every 3-6 months for 5 years after the initial surgery. ^12–15^

Circulating tumor DNA (ctDNA) has emerged as a promising noninvasive biomarker for molecular diagnosis of several cancer types.^16–22^ A pan-cancer analysis of ctDNA by Bettegowda et al. demonstrated that ctDNA detection rates were higher for CRC than for other cancer types.^23^ In CRC ctDNA was detectable in approximately 70% of patients with localized disease (stages I-III) and 100% of those patients with metastatic disease (stage IV).^23^ The clinical validity of ctDNA monitoring as defined by Merker et al.^24^ was demonstrated in terms of therapeutic efficacy in patients with metastatic CRC^25–28^ and for relapse prediction for those with localized CRC.^18, 21, 29, 30^ Recent prospective studies also reported that postoperative ctDNA status (i.e., positive or negative) can be used to stratify patients with resectable stage II/III CRC into patients who would likely benefit from adjuvant chemotherapy (ACT) and those for whom ACT can be safely omitted. ^31, 32^ Importantly, other investigators have suggested that reducing ACT doses according to ctDNA status realized substantial cost savings and improved quality-of-life.^33^

In daily practice, the most informative marker should quantitatively reflect the real time tumor burden rather than providing simple stratification based on snapshot molecular profiling. To carry out real time tumor burden monitoring using ctDNA, the marker should be sensitive, simple, and affordable, which are all essential factors for intensive surveillance. Although dPCR meets some of these requirements, preparation of a large number of validated mutation-specific primer/probe sets is a challenge in daily practice. In our previous work we overcame this challenge by establishing an original dPCR probe library including probes against >1,000 somatic mutations that are frequently found in human cancer, a resource we term Off-The-Shelf (OTS)-1000ex. The OTS-1000ex library allows immediate selection of validated dPCR primer/probe sets corresponding to somatic mutations identified in patient samples without need for optimization of dPCR conditions. Our previous studies also demonstrated that tumor-informed ctDNA monitoring has the clinical validity in terms of early relapse prediction, treatment efficacy evaluation, and non-relapse corroboration in management of several types of gastrointestinal cancers, including CRC.^34–37^ In the present study, we further investigated whether ctDNA monitoring by dPCR can alter intervals of CT testing during postoperative intensive surveillance without loss of critical therapeutic opportunities for clinical relapse.

## METHODS

### Patients and sample collection

This study was approved by the Institutional Review Board of Iwate Medical University (IRB #HGH28-15 and #MH2021-073). Written informed consent was obtained from all patients. Among 116 patients registered in the current study (UMIN Clinical Trial Registry: UMIN000045114), 52 who had undergone complete resection within at least 3 years of the initial surgery were enrolled (figure 1). These 52 patients were histologically confirmed to have CRC [n=4, 26, 20, and 2 having stage I, II, III, and IV disease, respectively] and were enrolled in the study between March 11, 2016, and June 20, 2018. All patients underwent primary tumor resection as a first-line therapy. Nearly all (51/52, 98.1%) of the patients underwent R0 resection during the initial operation. The remaining patient (CC16010) underwent two-stage resection with curative intent for the primary tumor and a metastatic liver tumor. Twelve patients (1 stage II, 10 stage III, and 1 stage IV) received ACT after curative resection. A summary of the patient characteristics is provided in online supplementary table S1. Surgically acquired primary tumor tissue samples and corresponding serial blood samples were obtained for somatic mutation screening and ctDNA monitoring, respectively.

**Figure 1.**
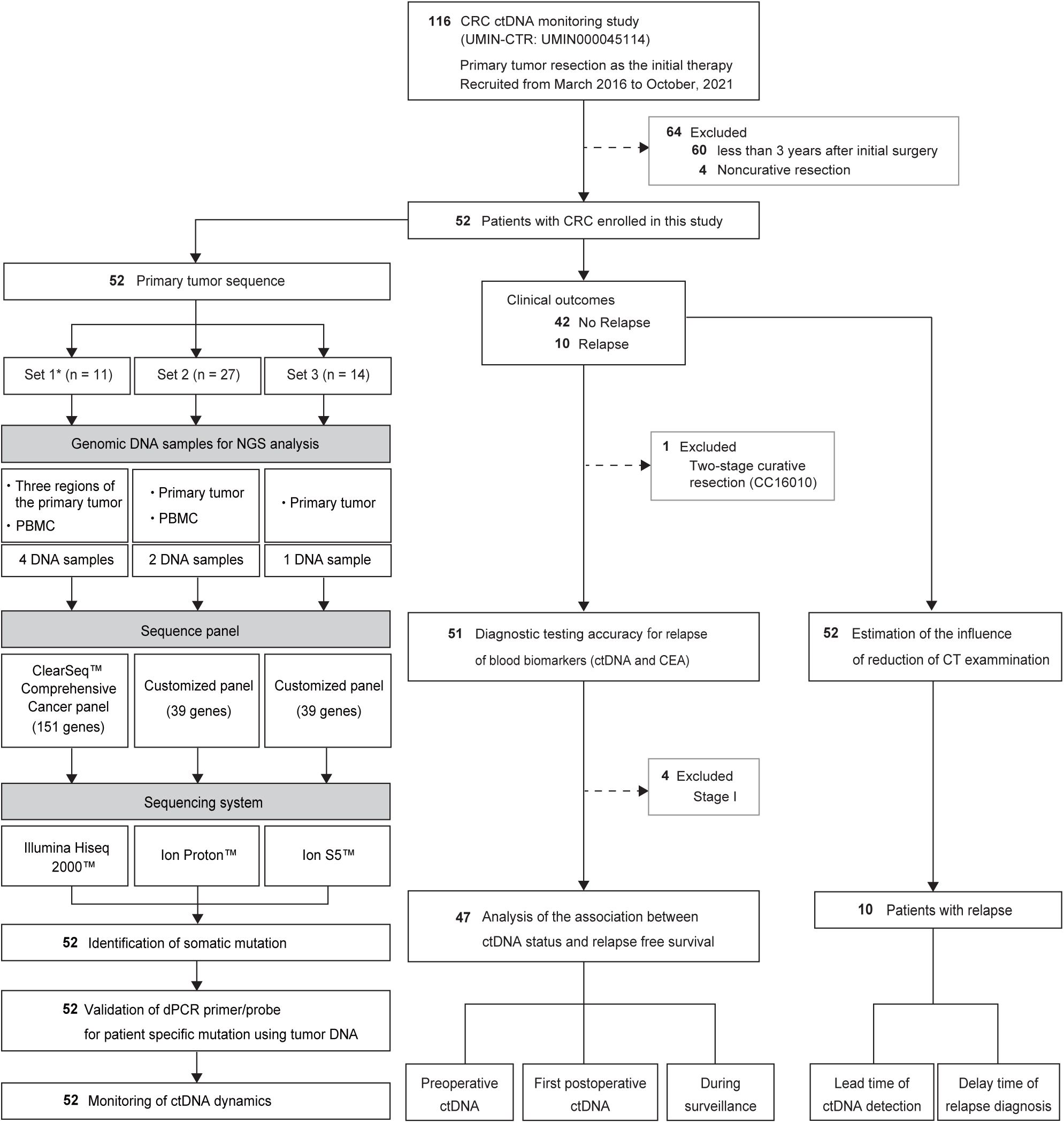
Flow of patient enrollment and sample collection during the study period. The selection of patients is shown with the type of platform used to analyze samples for the three study groups: Set 1, Set 2, and Set 3. CRC, colorectal cancer; ctDNA, circulating tumor DNA; UMIN-CTR, University Hospital Medical Information Network Center Clinical Trials Registry; NGS, next-generation sequencing; PBMC, peripheral blood mononuclear cell; dPCR, digital PCR; CT, computed tomography.

### Panel sequencing of primary tumor**s**

Samples were divided into three sets: Set 1, Set 2, and Set 3 (n=11, 27, and 14, respectively) and DNA samples from the tumor and corresponding peripheral blood mononuclear cells (PBMCs) were subjected to panel sequencing using three different platforms (online supplementary methods and figure 1). The highest priority of the primary tumor sequencing in this study was to detect a select number of somatic mutations that have high variant allele frequencies (VAF). Comprehensive descriptions of methods used to identify somatic mutations are presented in the online supplementary methods.^35, 37^

### Monitoring ctDNA levels using dPCR

The dPCR assay for quantitative monitoring of ctDNA levels was performed as described previously.^34–37^ Briefly, primers and probes labeled for wild-type and mutant alleles were specifically designed for each mutation identified in a primary tumor. Between 1 and 4 mutations per tumor that had a VAF >10% in primary tumors were prioritized for dPCR analysis. The criteria for mutation selection for the ctDNA assay and the definition of positive and negative ctDNA findings for dPCR are described in the online supplementary methods. ctDNA data for VAFs were plotted on a time course along with therapeutic regimens and clinical information. CEA levels were also measured at the same timepoints during ctDNA monitoring.

### Reduction of CTs for relapse detection during postoperative surveillance

In the relapsed group in the present cohort, patients underwent, on average, 3.3 CTs per year, whereas the number of CTs that actually returned a relapse diagnosis was 0.8 per patient per year. Hence, we hypothesized that reducing the number of CTs may have a limited effect on detection of relapse, particularly under circumstances in which ctDNA monitoring could complement relapse prediction by CTs. To test this hypothesis, we restricted the number of CTs timepoints to once per year and examined the relapse diagnosis. “Annual CT” timepoints were defined as CTs conducted at annual timepoints from periodic imaging examinations.

### Relapse detection using CT and ctDNA monitoring

The timing of clinical relapse was defined based on the timepoint at which a radiologist in daily clinical practice confirmed or suspected a lesion to represent a relapse (i.e., the word “relapse” was present in the radiographic report). Meanwhile, ctDNA relapse was defined as the time of the first ctDNA-positive detection of at least two consecutive ctDNA-positive timepoints. The lead time was defined as one day from the first ctDNA-positive indication of ctDNA relapse to the clinical relapse.

### Statistical analysis

For contingency table analysis, diagnostic ability to detect clinical relapse by both ctDNA and CEA was expressed as sensitivity/specificity, as well as positive/negative predictive values. Over the course of serial monitoring, ctDNA was defined as positive when the VAF was above the below detection limit (BDL). CEA values were binarized according to the upper limit of the normal level. For group comparisons, Mann-Whitney U and Fisher’s exact tests were used. Kaplan-Meier estimates with log-rank tests were used to compare relapse-free survival (RFS) and were stratified based on the ctDNA status (i.e., positive or negative) before treatment, at the first postoperative timepoint after the initial surgery, and throughout the postoperative surveillance period. Clinical RFS based on CT findings, estimated clinical RFS based on annual CT, and ctDNA-RFS based on ctDNA status were also compared. A Cox proportional hazards model was used to estimate risks. *P* values <0.05 were considered to be statistically significant for all analyses. All analyses were performed using GraphPad Prism 8 (GraphPad Software, San Diego, CA).

## RESULTS

### Mutations in primary CRC

At least one somatic mutation was identified in all 52 patients. The samples were divided into three sets, Set 1, Set 2 and Set 3. Online supplementary tables S2 and S3 and figure S1 summarize the mutation profiles. For Set 1, the detailed mutation profile is available in our previous report.^35^ For Set 2, an average of 3 mutations per sample (range, 1-24) and for Set 3 an average of 3 mutations per sample (range, 1-36) were identified using the respective customized panels. Among the 3 different sequencing platforms, the most frequently mutated genes were *TP53* (37/52, 71.2%), *APC* (28/52, 53.8%), *KRAS* (24/52, 46.2%), *PIK3CA* (13/52, 25.0%), and *BRAF* (8/52, 15.4%). The average VAFs for these five genes were: *TP53*, 44.8% (range, 7.78%-85.9%), *APC*, 33.3% (range, 12.0%-61.0%), *KRAS*, 31.1% (range, 3.6%-81.7%), *PIK3CA*, 28.5% (range, 8.0%-40.5%), and *BRAF*, 28.1% (range, 12.9%-48.2%). Among the 52 patients, 50 (96.2%) had mutations in at least one of the five genes, and 41 of 52 (78.8%) had more than two mutations in one of the five genes.

### Mutations selected for ctDNA detection

A total of 87 mutations were selected from the 52 patient samples according to our mutation selection algorithm (online supplementary methods) for dPCR. On a per-patient basis, the number of mutations used for ctDNA testing ranged between 1 and 5 (1: 30 patients; 2: 13 patients; 3: 6 patients; 4: 2 patients and 5: 1 patient (mean 1.7 ± 0.8)). Among the 87 mutations analyzed by dPCR, 48 (55.2%) were recurrent and covered by 14 primer/probe sets (figure 2). Preoperative plasma from 31 of the 52 (59.6%) patients was positive for ctDNA (figure 3). Of these preoperative ctDNA-positive patients, 1/4 (25.0%), 14/26 (53.8%), 15/20 (75.0%), and 1/2 (50%) were stage I, II, III, and IV disease, respectively.

**Figure 2.**
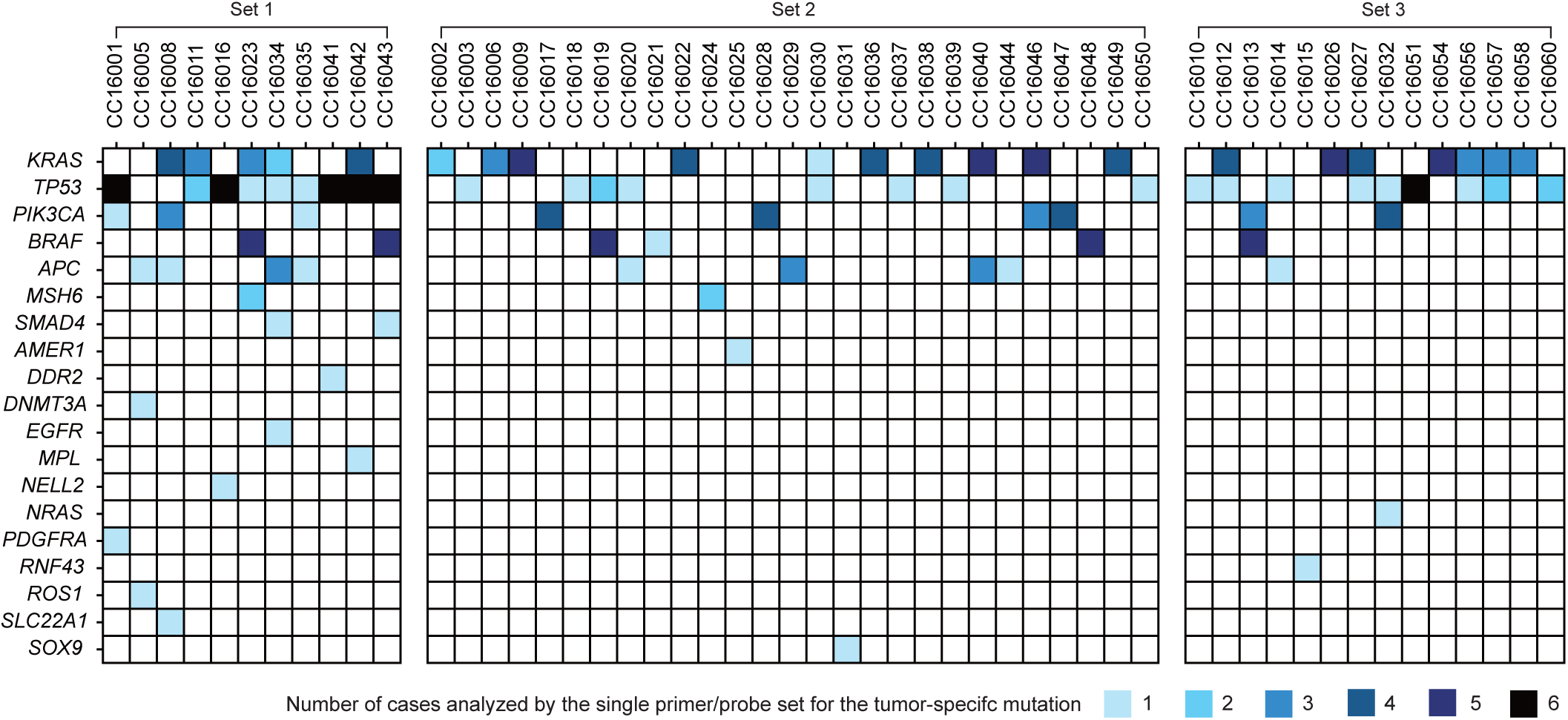
Somatic mutation profile of primary CRC tumors analyzed by dPCR. Mutation profile of the 52 CRC tumors analyzed by dPCR is shown. Colored boxes indicate the intersection between patients and primer/probe sets. Different shades of blue indicate the number of cases analyzed using a single primer/probe set for the indicated tumor-specific mutation. Patient number is arrayed along the top and each row corresponds to a different gene.

**Figure 3.**
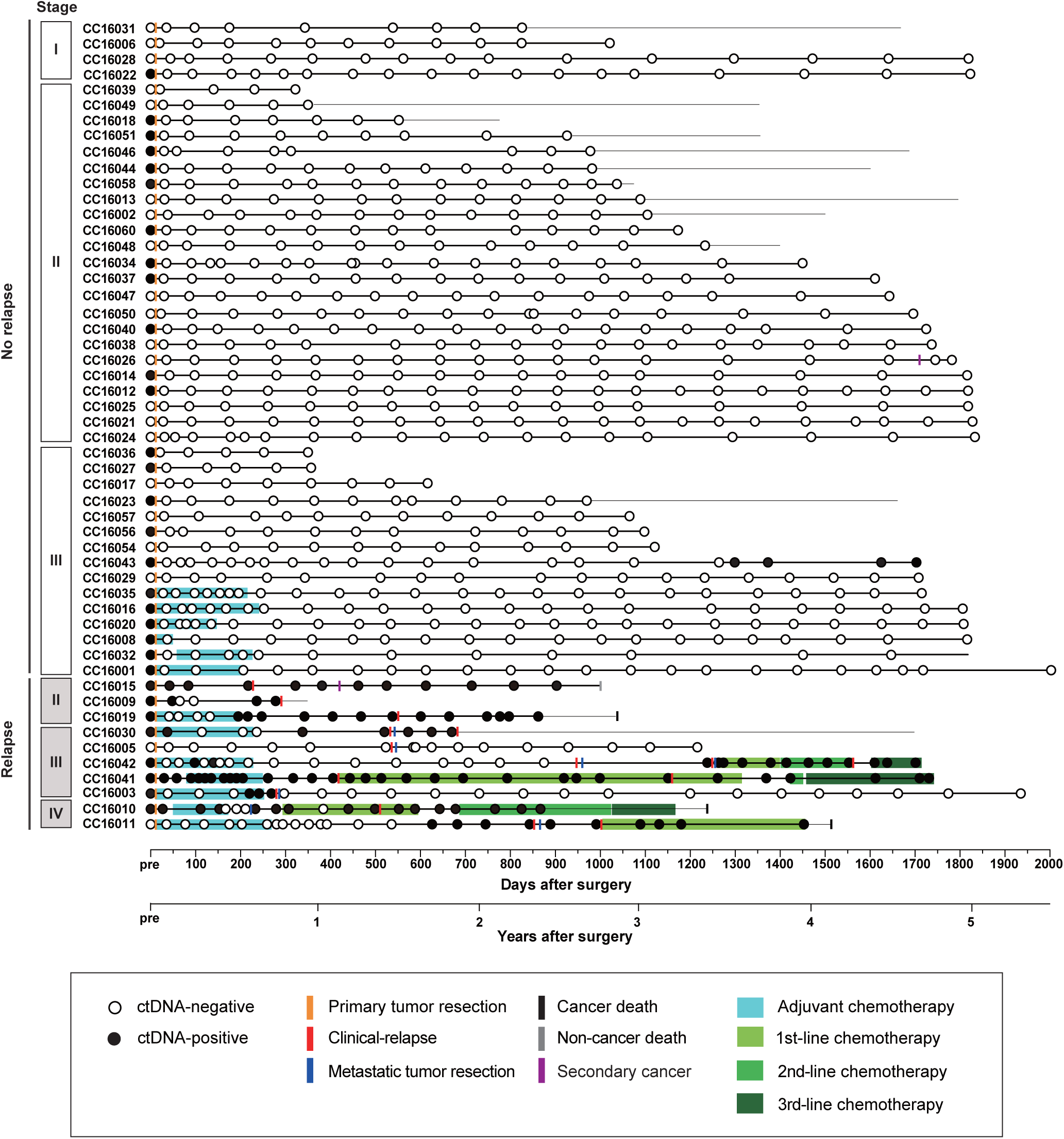
Longitudinal ctDNA monitoring during the postoperative period for patients with CRC. Patient numbers are grouped according to disease stage and relapse status. The days after initial resection surgery are shown on the bottom. Open and filled circles indicate ctDNA-negative and -positive, respectively. Orange bars designate date of primary tumor resection. Red bars denote date of clinical relapse and blue bars show day of surgery for resection of metastatic tumors. Black, gray, and purple bars denote day on which death from cancer, non-cancer cause, or secondary cancer, respectively, occurred. Duration of adjuvant (teal) and 1^st^ (light green)-, 2^nd^ (green)- and 3^rd^ (dark green)-line chemotherapy is indicated. ctDNA, circulating tumor DNA. Horizontal bars indicate observation periods.

### Longitudinal ctDNA monitoring in postoperative surveillance

The median observation period was 1,503 days (range, 322-1,951 days). A total of 1,526 plasma samples from 867 timepoints, resulting in 16.7 analyzable timepoints per patient, were analyzed for ctDNA. The first postoperative plasma samples from the initial surgery were collected an average of 34.0 days (range, 20-58 days) after resection. To categorize longitudinal data, ctDNA levels for each timepoint were binarized (i.e., ctDNA-positive and -negative; figure 3). Clinical relapse occurred for 10 out of the 52 (19.2%) patients with CRC. Patients who had clinical relapse had significantly higher ctDNA-positive rates at all analyzed points compared to patients who did not have relapse [59.1% (114/193) vs. 4.0% (27/674), *P* <0.0001, Fisher’s exact test]. The ctDNA dynamics and detailed clinical information for the 10 relapsed and 42 non-relapsed patients are shown in figure 4 and online supplementary figure S2, respectively. For the 42 patients without relapse, ctDNA-negative results were obtained throughout the postoperative surveillance period, which ranged from 10.7 months to 65.0 months. Of these 42 non-relapsed patients, 23 were preoperative ctDNA-positive. However, these 23 patients exhibited a ctDNA level that was below the detection limit at the first postoperative timepoint after the initial surgery and they continued to be ctDNA-negative. Meanwhile, for the 10 patients that had clinical relapse, 9 (90%) had an increased ctDNA level when clinical relapse and tumor growth was noted; the ctDNA level decreased in these patients in response to treatment for the relapse. Taken together, these results indicated that ctDNA monitoring by dPCR provided valid information for tumor burden estimation during the clinical course for nearly all enrolled patients (51/52; 98.7%).

**Figure 4.**
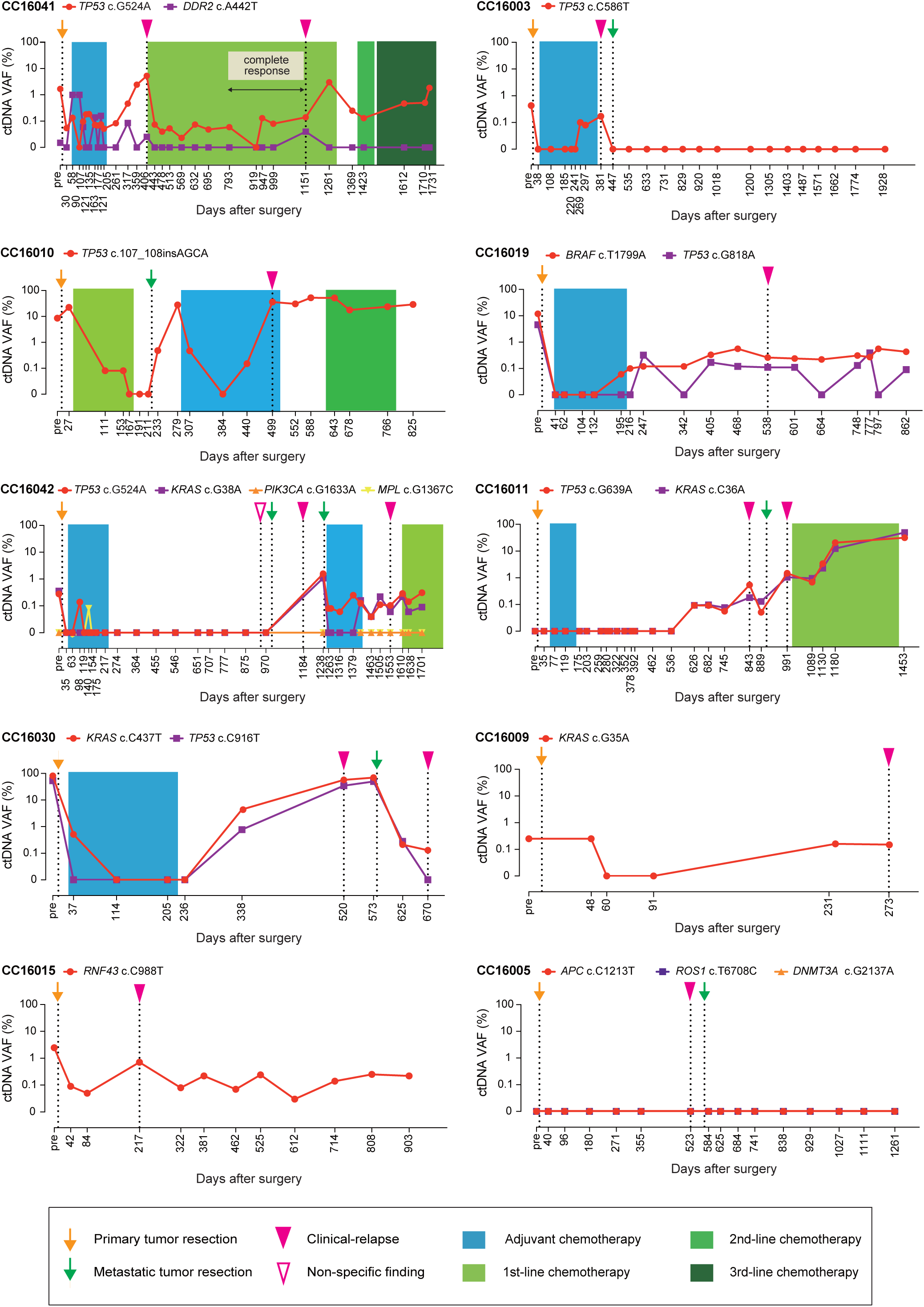
Dynamics of ctDNA during the postoperative period for CRC patients with relapse. The history of 10 patients who experienced relapse is shown. The days post-surgery are on the x-axis and the percentage ctDNA VAF is on the y-axis. Teal and green rectangles indicate duration of chemotherapy regimens. Genes carrying mutations are shown beside the patient number and the colors of the genes correspond to the plot of VAF values. Orange and green arrows indicate time of primary and metastatic tumor resection, respectively. Open and filled pink triangles note the day of a non-specific finding or determination of clinical-relapse, respectively. CRC, colorectal cancer; ctDNA, circulating tumor DNA; VAF, variant allele frequency.

### Patterns of ctDNA dynamics in a therapeutic context

The dynamics of ctDNA during the postoperative surveillance period in patients with clinical relapse who received ACT could be divided into 3 patterns: continuously positive (Patient CC16041); ctDNA elevation during ACT (Patients CC16003, CC16010, CC16019 and CC16042); and ctDNA elevation post-ACT (Patients CC16011 and CC16030; figure 4). In the surgery-only patients who had clinical relapse, the ctDNA dynamics had either fluctuations in ctDNA positivity (Patients CC16009 and CC16015) or were continuously ctDNA-negative (Patient CC16005) (figure 4). Serial ctDNA analysis identified clinical relapse with 85.7% sensitivity and 97.6% specificity as well as a 92.3% positive predictive rate (PPR) and 95.4% negative predictive rate (NPR) during the postoperative surveillance period.

### CEA levels in the context of clinical relapse

The CEA level for each timepoint was also binarized (i.e., above and below the upper limit of normal) in a swimmer plot (online supplementary figure S3). Unlike ctDNA, positive CEA levels were often observed during the postoperative period in patients who did not have clinical relapse (120/613 [19.6%] timepoints; 16/42 [38.1%] patients). Furthermore, for clinical relapse patients the dynamics of CEA levels did not reflect changes in tumor burden as accurately as ctDNA levels did. The diagnostic performance of serial CEA analysis for clinical relapse was 64.3% sensitivity, 61.9% specificity and 36.0% PPR, and 83.9% NPR.

### Plasma ctDNA status and risk of recurrence

We evaluated, excluding Stage I patients (n=4), the 5-year RFS rate stratified with ctDNA status for 47 patients. There was no significant difference in 5-year RFS between preoperative ctDNA-positive (n=28) and -negative (n=19) groups (HR 2.5, 95% CI 0.7–9.4, *P=*0.22, log-rank test; figure 5A). Patients who had a ctDNA-positive finding at the first postoperative timepoint after the initial surgery (n=4) showed a significantly higher risk of clinical relapse than those who were ctDNA-negative (n=43) (HR 39.6 (95% CI: 6.4–243.9) *P* < 0.0001, log-rank; figure 5B). Similarly, patients who had at least one ctDNA-positive timepoint during the postoperative surveillance period (n=9) showed a significantly higher risk of clinical relapse than those who had sustained ctDNA-negative results (n=38) (HR 56.3, 95%CI 7.8-407.0, *P* < 0.0001, log-rank test; figure 5C).

**Figure 5.**
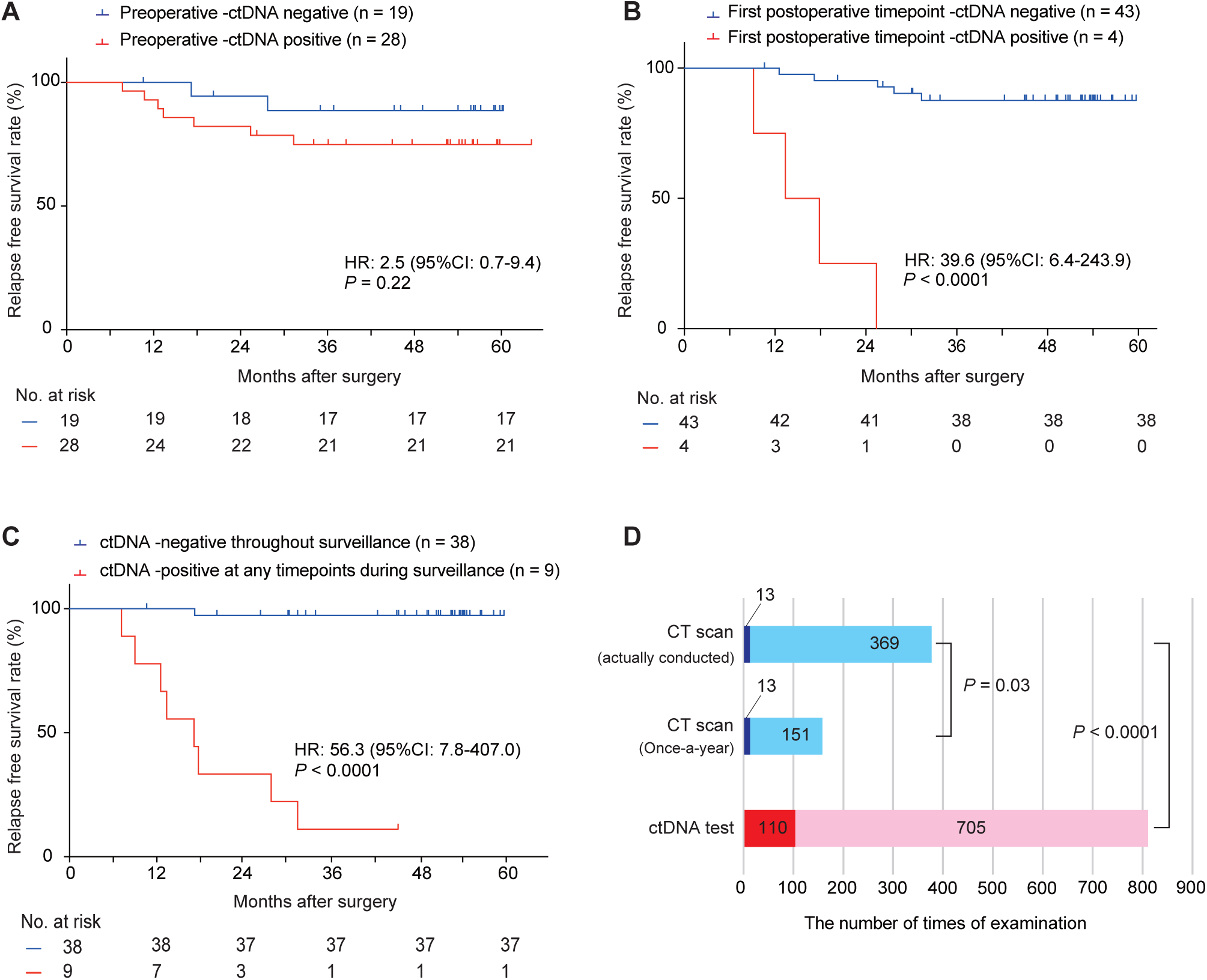
Association between ctDNA status and clinical relapse. (A)–(C) Relapse-free survival according to ctDNA status at (A) Preoperative timepoint; (B) First postoperative timepoint after initial surgery and (C) During the postoperative surveillance period. HR, hazard ratio. *P* values were derived from a Kaplan-Meier log-rank test. HR was calculated using the log-rank test. (D) The number of CT and ctDNA assays during the postoperative surveillance period. Dark and light blue boxes indicate the number of CTs with or without confirmation of clinical relapse, respectively. Red and pink boxes indicate the number of ctDNA-positive and -negative results, respectively. CT, computed tomography scan; ctDNA, circulating tumor DNA.

### Possibly unnecessary CTs during postoperative surveillance

Patients in the study cohort underwent 382 CT procedures, with an average frequency of 3.3/year per patient. Of these CTs, 13 (3.4%) procedures contributed to delivery of a relapse diagnosis (including multiple relapses per patient) (figure 5D). The timing of the CT during the postoperative periodic surveillance period for all patients is presented (figure 6 and online supplementary figure S2). If an annual interval for CT procedures was used during the surveillance period, fewer CTs would have been carried out (164 vs. actual 382). With an annual interval, the relapse detection rate per CT increased from 3.4% (13/382) to 7.9% (13/164), meaning that over half the CTs performed might have been unnecessary in terms of relapse detection (figure 5D). By extending periodic CT intervals to an annual basis, potential delays in detection of clinical relapse detection might have occurred for 7 of 10 patients in 9 of 13 events, with a median delay of 164 days (range, 0-267; figure 6). Hence, extending the interval to an annual basis is not recommended if clinical relapse detection relies only on CT.

**Figure 6.**
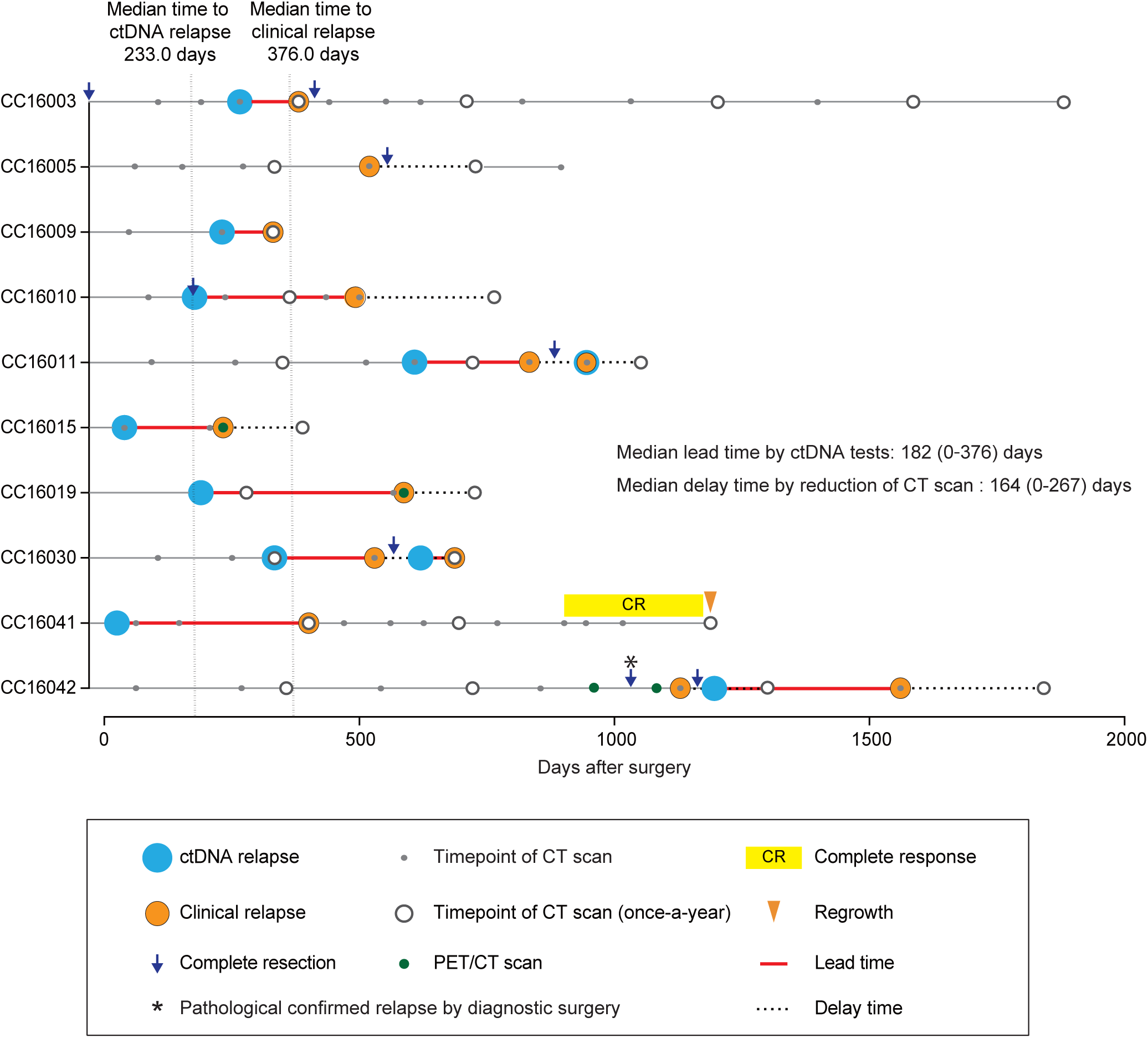
Lead time for ctDNA elevation and delay in relapse detection during the surveillance period with CT performed annually to detect clinical relapse in CRC patients. All time points for CT (gray dots and open circles) and the first time points for ctDNA detection after curative resection during the relapse-free period are shown. Blue circles indicate date of ctDNA relapse and orange circles indicate date of clinical relapse. Red lines correspond to lead time between ctDNA relapse detection and clinical relapse. Dotted lines indicate delay time in detection of relapse by annual CT. CRC, colorectal cancer; ctDNA, circulating tumor DNA; CT, computed tomography scan; PET, positron emission tomography.

In contrast, the fraction (13.5%, 110/815) of ctDNA-positive timepoints among all timepoints was significantly higher than the fraction of relapse-confirming CTs among all CTs carried out, suggesting that ctDNA may have more rigorous detection ability than CTs (3.4 %, *P* < 0.0001, Fisher’s exact test, figure 5D). A ctDNA elevation (i.e., ctDNA-relapse) was observed before the actual clinical relapse (11 of 13 events) for 9 of the 10 clinical relapse patients. The median lead time for this elevation is 182 days (range, 0-376 days; figure 6), which is largely consistent with previous studies (5.6-11.5 months).^18, 21, 30^ If the CT procedure was set for an annual interval during surveillance, the estimated lead time from the ctDNA relapse to the clinical relapse with annual CT was extended to 339 days (range, 42-533 days).

Currently, RFS is largely dependent on the timing of CT as most relapses are diagnosed by periodic CT conducted every 3-4 months. Hence, we next evaluated how increasing the interval of periodic CT (i.e., conventional CT) to annual (i.e., annual CT) could affect RFS. Here ctDNA-RFS was calculated with the event, “relapse”, defined as the first day ctDNA elevation was detected. We found 9 patients who had analyzable events that could be defined as both clinical- and ctDNA-relapse. No significant difference in the clinical-RFS rate was seen based on CT interval (i.e., conventional vs. annual CT (online supplementary figure S4A)). In contrast, the conventional clinical RFS rate was significantly higher than that for ctDNA RFS (online supplementary figure S4B). Furthermore, a larger difference in recurrence rates was seen between annual clinical RFS and ctDNA RFS (online supplementary figure S4C).

## DISCUSSION

Longitudinal ctDNA monitoring during postoperative surveillance reportedly can predict clinical relapse in patients with CRC.^18, 21, 30^ Although the patient cohort in the present study was of moderate size (n=52), here we prioritized detailed (i.e., 16.7 timepoints per patient) and long-term ctDNA monitoring (i.e., median observation time 50.0 months). The dPCR system that we developed using OTS-Probes also allows essential ctDNA monitoring quality and is supported by the finding that patients who had ctDNA-positive results had significantly higher risk for clinical relapse than those who were ctDNA negative. In the present study, in addition to the longitudinal ctDNA monitoring, we combined radiographic and clinical data to evaluate whether ctDNA monitoring could compensate for a reduction in the frequency of periodic CTs without compromising the opportunity for relapse detection during postoperative surveillance.

One challenge associated with extending CT intervals during disease surveillance is that the likelihood of a delay in relapse detection could increase. Conventionally, intensive surveillance involving a CT every 3 months would allow early detection of recurrence after surgical resection for colorectal metastasis.^12, 38^ Indeed, patients in this study underwent CT on average 3.3 times/year. In patients who had clinical relapse, small lesions could often be retrospectively identified with a CT several months before a diagnosis of clinical relapse was returned. However, suspicious, but marginal, findings by CT alone did not result in timely treatment interventions for relapse. In terms of the timing of the clinical relapse diagnosis, in the present study we saw no significant difference in clinical RFS rates between conventional and annual CT intervals. While these results indicate that an annual CT interval would be acceptable in terms of relapse detection, additional modalities like ctDNA monitoring may be needed to improve outcomes achieved with timely therapeutic intervention.

CEA is one of the most widely used serum tumor markers to measure tumor burden in postoperative CRC surveillance.^39^ In fact, a previous study showed that CRC patients who had elevated postoperative CEA had an increased risk of recurrence.^40^ In the present study, our results suggest that ctDNA monitoring reflects tumor burden more accurately than CEA (50/52, 96.2% vs. 33/52, 63.5%). When ctDNA was used as tumor marker, the median lead time from ctDNA relapse to clinical relapse was 182 days (range, 0-376 days) suggesting that tumor-specific ctDNA detection may indeed be helpful for relapse diagnosis of small lesions that are classified as marginal findings or cannot be detected with a CT. If surveillance with annual CTs and periodic ctDNA monitoring is carried out, diagnosis of clinical relapse may be delayed, whereas a ctDNA relapse diagnosis will be delivered in more timely manner. Although the “ctDNA-RFS rate” would be smaller than the “clinical-RFS rate”, patients could have more opportunities for therapeutic intervention at an early stage of relapse. Overall, our findings support a rationale in which CTs for relapse detection that are conducted at longer intervals would not delay relapse diagnosis if compensatory data is provided by frequent ctDNA monitoring (e.g., every 3 months).

In terms of ACT for patients with resectable CRC, recent prospective studies demonstrated that a ctDNA-guided approach based on ctDNA status determined 4 or 7 weeks after surgery could reduce ACT use and identify patients who are more likely to benefit from ACT.^31 32^ Our present dPCR-based ctDNA monitoring also demonstrated that all 42 patients without relapse were ctDNA-negative at the first postoperative timepoint that occurred between 3 and 7 weeks after surgery (figure 3). However, 5 of 10 (50.0%) patients with clinical relapse were also ctDNA-negative up to 8 weeks after surgery (figure 4). These ctDNA-negative clinical relapse patients may not have received ACT if a ctDNA-guided approach was used for therapeutic decision making, and thus could have had early relapse. Therefore, some patients may not benefit from elimination of ACT based on ctDNA-negative results returned at only one or two postoperative timepoints. In addition, reports of ctDNA-guided therapy used ultra-deep sequencing for qualitative judgement.^31^ ^32^ As the principle of current NGS includes technological limitations for stable quantification of very low VAF samples such as ctDNA from blood, this approach may be difficult to use for serial monitoring of ctDNA.^41^ However, as we demonstrated by measurements using dPCR, the dynamics of <1% ctDNA VAFs reflect clinically important information. The ability to detect VAFs as low as 0.001% by dPCR^42^ suggests that a ctDNA-negative result is strongly indicative of disease-free status. Our frequent ctDNA monitoring in the present study revealed that the median lead time from ctDNA relapse to clinical relapse was 6 months, which in fact was indicated by less than 1% of serial VAFs. Therefore, a ctDNA-negative status at the first postoperative timepoint does not necessarily indicate a lack of benefit from ACT. Instead, our results indicated that frequent ctDNA monitoring may be necessary to guide and confirm optimal personalized treatment opportunities.

The current study has some potential limitations. First, most of the serial ctDNA monitoring involved samples with 1 or 2 mutations. Although this number of mutations seems to be sufficient to reflect tumor burden during the surveillance period, it does not fully detect changes in peripheral genetic heterogeneity during therapies. To monitor such heterogeneity, panel sequencing is required although NGS may not be suitable for monitoring of tumor genetic heterogeneity with extremely low quantification of VAF ctDNA.^41^ Second, therapeutic regimens were not selected based on common criteria. Companion diagnostics were occasionally used for anti-EGFR antibody therapy whereas chemotherapeutic regimens were selected according to guidelines, as well as doctor/patient preference. Finally, although the median follow-up time of the present study is over four years, whether long-term follow-up for relapse is warranted is unclear.

In conclusion, ctDNA monitoring can extend the interval for CT from every 3-4 months to annually during the postoperative surveillance period for CRC patients without increasing the likelihood that relapse diagnosis would be delayed. To accomplish this quality of results, ctDNA monitoring should be sensitive (i.e., 0.1% VAF detectable), frequent (i.e., every 3-4 months) and use a suitable number of validated probes against personalized somatic mutations in CRC. This level of frequency should be attainable using an affordable technology such as dPCR.

## Data Availability

All data produced in the present work are contained in the manuscript.

## Acknowledgments

Author contributions: Takeshi Iwaya, Tomoko Sasaki, Mizunori Yaegashi, and Satoshi S. Nishizuka had full access to all study data. They take responsibility for the integrity of the data and the accuracy of the data analysis. Acquisition, analysis, or interpretation of data: Takeshi Iwaya, Satoshi S. Nishizuka, Tomoko Sasaki, Mizunori Yaegashi, Yuka Koizumi, Akiko Yashima-Abo, Noriyuki Sasaki, Ryosuke Fujisawa, Fumitaka Endo, Shoichiro Tange, Tomomi Hirano, Masashi Idogawa, Yasushi Sasaki, Masashi Fujita, and Hidewaki Nakagawa. Drafting of the manuscript: Tomoko Sasaki, and Takeshi Iwaya. Critical revision of the manuscript for important intellectual content: Hayato Hiraki, Masakazu Abe, Yasushi Sasaki, Takashi Tokino, and Satoshi S. Nishizuka. Administrative, technical, or material support: Mizunori Yaegashi, Koki Otsuka, Akira Sasaki, Mari Masuda. Study supervision: Satoshi S. Nishizuka, Yasushi Sasaki, Mari Masuda, Fumiaki Takahashi, and Takashi Tokino. We thank the surgeons in the Department of Surgery, Iwate Medical University, Drs. Toshimoto Kimura, and Kiyoharu Takashimizu.

## Conflict of Interest

Dr. Iwaya received grant/research support from Nippon Kayaku, Chugai Pharmaceutical, Daiichi Sankyo and Quantdetect Inc. Dr. Iwaya is a consultant of Quantdetect Inc. Drs. Hiraki and Nishizuka received financial/research support from LSI Medience Co. and Quantdetect Inc. Dr. Hiraki is a consultant of Quantdetect Inc. Dr. Nishizuka received grant/research support from Taiho Pharmaceuticals, Boehringer-Ingelheim, Chugai Pharmaceutical, and Geninus. Dr. Nishizuka received grant/honoraria from Thermo Fisher Scientific, MSD, LSI Medience, and Finngal Link. Dr. Nishizuka is an advisor of CLEA Japan and Hitachi High-Tech Co. Dr. Nishizuka is Founder and CEO of Quantdetect Inc. Drs. Iwaya and Nishizuka hold a patent that might benefit from this publication (JP6544783).

## Funding

This work was supported by a Grant-in-Aid for Scientific Research KAKENHI [16K19951, 16K19952, 16H06279, 17K10605, 19K09224, 20K09064, 20K08966, and 21K07223], Keiryokai Collaborative Research Grant [#136 and #145], Iwate Prefectural Research Grants [H30, R2, and R3], and AMED under Grant number JP23ck0106825.

## Notes

### Competing Interest Statement

The authors have declared no competing interest.

### Clinical Trial

UMIN Clinical Trial Registry number, UMIN000045114

### Author Declarations

This study was approved by the Institutional Review Board of Iwate Medical University (IRB #HGH28-15 and #MH2021-073).

### Summary of Updates

Title and content with unclear descriptions have been revised to make the research purpose, results, and discussion clear. Authors have removed unessential figures in previous version (i.e., Figure 2B, 2C, 5A, 5B, Supplementary Figure 2, 3, 6, 7, and 9).

