## Supplementary File for "Impact of sensitive circulating tumor DNA monitoring on CT scan intervals during postoperative colorectal cancer surveillance"

### Supplementary Material

**Supplementary Methods**

**Supplementary Table S1.** Clinicopathological characteristics of patients in the current study cohort.

**Supplementary Table S2.** Primary tumor mutations detected through sequence analysis of Set 2 samples using Ion Proton™ and circulating tumor DNA levels (ctDNA) measured by digital PCR.

**Supplementary Table S3.** Primary tumor mutations detected through sequence analysis of Set 3 samples using Ion 5S™ and circulating tumor DNA (ctDNA) levels measured by digital PCR.

**Supplementary Figure S1.** Somatic mutation profile of primary CRC tumors from the 52 patients in the current study cohort.

**Supplementary Figure S2.** Dynamics of ctDNA in 42 patients with CRC without clinical relapse.

**Supplementary Figure S3.** Longitudinal CEA monitoring of patients with CRC during the postoperative period.

**Supplementary Figure S4.** Comparison of relapse free survival (RFS) rates according to relapse type.

#### Supplementary Methods

##### Identifying somatic mutations in primary tumor tissue

Sequencing analysis of the primary tumor was performed using three different platforms for three sets of patients with colorectal cancer (CRC). Tumor samples from 11, 27, and 14 patients were analyzed for Sets 1, 2, and 3, respectively. Set 1 was analyzed using an Illumina Hiseq 2000 sequencer (Illumina, Inc., San Diego, CA) with the ClearSeq Comprehensive Cancer Panel (Agilent Technologies, Inc., Santa Clara, CA) that targets 151 disease-associated genes (Ref. 35 of the main text). Sets 2 and 3 were analyzed using the Ion Proton™ and Ion S5™ system (Thermo Fisher Scientific, Waltham, MA), respectively, with a customized panel targeting 39 genes that are frequently altered in CRC.[1] Set 1 samples were analyzed in our previous study, which considered three regions of the primary tumor and corresponding DNA from peripheral blood mononuclear cells (PBMCs) (total of four samples) to assess the effect of intra-tumor genetic heterogeneity on circulating tumor DNA (ctDNA) (Ref. 35 of the main text). In that study, the results indicated that tumor genetic heterogeneity did not present a major obstacle for ctDNA analysis provided that a somatic mutation having a sufficient variant allele frequency (VAF) was selected from a single region of the tumor. Furthermore, commonly detected mutations among the three tumor regions were limited to a set of genes that included *TP53*, *APC*, *KRAS*, *PIK3CA*, *FBXW7*, and *BRAF*.

For Set 2, the pairing between DNAs from tumor andPBMC, was analyzed using an Ion Proton™ system (Thermo Fisher Scientific, Waltham, MA) with a customized panel targeting 39 genes specific for CRC. This approach was designed to reduce costs and improve the efficiency of mutation detection by tumor sequencing. Amplicon sequencing followed by identification of somatic mutations using the tumor-normal workflow of Ion Reporter software 5.0 (Thermo Fisher Scientific) was performed as previously described (Refs. 34 and 37 of the main text). The following criteria were used as cutoffs: (1) total coverage >200, (2) variant coverage >10, and (3) variant frequency >10%. Mutations were called if they occurred in <0.1% of the reads in the normal control (minor allele frequency) and were absent from the dbSNP and 1000 Genomes Project database.

For Set 3, we analyzed only tumor DNA samples from a single region using an Ion S5™ system (Thermo Fisher Scientific). This approach considered the high mutation detection efficiency of tumor sequencing using the customized panel for Set 2. Alignment to reference genomes and sequencing read counting were performed with Torrent Suite version 5.0 (Thermo Fisher Scientific), Hisat2 (https://ccb.jhu.edu/software/hisat2/index.shtml), and BWA (Burrows–Wheeler

Alignment tool). For such cases, commonly-detected mutations among the three algorithms were prioritized in subsequent mutation selection. The following criteria were used as cutoffs: (1) total coverage >200, (2) variant coverage >10, and (3) variant frequency >10%. Mutations were selected as described for Set 2. Furthermore, mutations were called if they also occurred with minor allele frequencies (MAF <0.01) in the Japanese SNP database of the Tohoku Medical Megabank Organization (ToMMo).[2] Moreover, the same mutations shared among more than 3 of the 14 patients in Set 3 were excluded from selection since the mutations did not frequently occur in CRC and thus the likelihood that they resulted from sequencing errors was high. For insertion and deletion mutations, the following criteria were used as cutoffs: (1) total coverage > 200; (2) variant coverage >10; and (3) variant frequency >20%. For Set 3, the dPCR probe/premier set for each specific mutation was designed after confirming that Sanger sequencing detected the mutation in tumor DNA but not in PBMC DNA.

Clonal hematopoiesis of indeterminate potential variants was removed by paired PBMC sequencing for Sets 1 and 2, and for Set 3 by confirming the absence of the corresponding specific mutation in PBMC DNA by Sanger sequencing.

**Criteria for mutation selection in ctDNA assays using digital PCR**

Specific primers and probes labeled for wild-type and mutant alleles were specifically designed for each mutation identified in a primary tumor using Hypercool Primer & Probe™ technology (Nihon Gene Research Laboratories, Sendai, Japan). For frequently recurring missense mutations, commercially available primer/probe sets were used (Quantdetect, Inc., Tokyo, Japan; Thermo Fisher Scientific, Waltham, MA). According to our previous studies, between 1 and 5 mutations having VAFs >10% that were found in primary tumors were prioritized for digital PCR (dPCR) analysis (References 34-37 of the main text). The criteria used to select mutations for ctDNA assay were: (1) the mutation having the highest VAF in the primary tumor; (2) up to four additional mutations having a VAF >10% in the primary tumors; (3) recurrent mutations observed among patients; (4) mutated genes having a high success rate for primer/probe validation; and (5) exclusion of mutations with a surrounding sequence that was suspected of having low PCR efficiency and low specificity of hybridized probes (e.g., self-complementarity and/or secondary structure of the primer/probe). Furthermore, those mutations that were already validated in tumor DNA from CRC or other types of malignancies could reduce the temporary cost and time needed for syntheses of specifically-designed probes for each tumor-specific mutation. Therefore, mutations listed in our dPCR primer/probe library and commercially available primer/probe sets

(Quantdetect, Inc, Tokyo, Japan) were analyzed, even if the mutations detected in the primary tumors did not satisfy the criteria for mutation selection.

**Definitions of ctDNA positivity and negativity in dPCR assays**

As previously described (References 34-37 of the main text), dPCR was performed using the QuantStudio3D Digital PCR system (Thermo Fisher Scientific). After PCR amplification, chips were read on a QuantStudio 3D Digital PCR Instrument (Thermo Fisher Scientific), and a secondary analysis was performed using QuantStudio 3D Analysis Suit Cloud software (Thermo Fisher Scientific). For further quality control, the boundaries of FAM, VIC, and undetermined events were manually defined. After confirmation of sufficiently separate reactions of both wild-type and mutant alleles in tumor DNA, plasma DNA was evaluated by dPCR. Plasma samples having more than two positive reactions (i.e., dots) for mutant DNA were defined as ctDNA-positive. If only one positive reaction for mutant DNA was detected, samples were considered to be ctDNA-positive when more than one positive reaction for mutant DNA could be repeatedly detected upon retesting of the same sample. The VAF values were calculated as the fraction of mutant/(mutant + wild) divided by the number of partitions in which either the mutant or wild-type sequence was detected.[3] If no mutant partitions were detected, the case DNA was regarded as "negative".

**Supplementary Table S1.**
**Clinicopathological characteristics of patients in the current study cohort.**

| Patient No. | Age range | Sex | Primary tumor location | Size (mm$^2$) | UICC stage [a] | | | | | Preoperative metastatic organ |
|---|---|---|---|---|---|---|---|---|---|---|
| | | | | | T | depth | N | M | pStage [b] | |
| CC16001 | 71-75 | Female | Cecum | 47×60 | 4a | SE | 1a | 0 | IIIB | − |
| CC16002 | 81-85 | Female | Cecum | 57×35 | 3 | SS | 0 | 0 | II | − |
| CC16003 | 51-55 | Female | Rectum | 23×22 | 3 | SS | 2 | 0 | IIIB | − |
| CC16005 | 71-75 | Female | Rectosigmoid | 22×20 | 2 | MP | 1b | 0 | IIIA | − |
| CC16006 | 51-55 | Male | Sigmoid | 9×11 | 1b | SM | 0 | 0 | I | − |
| CC16008 | 71-75 | Female | Cecum | 26×20 | 2 | MP | 1b | 0 | IIIA | − |
| CC16009 | 86-90 | Female | Rectum | 60×75 | 4a | SE | 0 | 0 | II | − |
| CC16010 | 56-60 | Female | Rectum | 35×70 | 4a | AI | 2b | 1a | IVB | Liver |
| CC16011 | 51-50 | Female | Sigmoid | 37×48 | 4a | SE | 1a | 1a | IVA | LN [c] |
| CC16012 | 56-60 | Female | Sigmoid | 53×49 | 3 | SS | 0 | 0 | II | − |
| CC16013 | 76-80 | Female | Rectum | 65×50 | 3 | SS | 0 | 0 | II | − |
| CC16014 | 51-55 | Female | Rectum | 85×57 | 3 | SS | 0 | 0 | II | − |
| CC16015 | 71-75 | Male | Sigmoid | 9×11 | 4b | SM | 0 | 0 | IIC | − |
| CC16016 | 51-55 | Male | Sigmoid | 28×28 | 3 | SS | 1a | 0 | IIIB | − |
| CC16017 | 76-80 | Female | Ascending | 103×72 | 3 | SS | 1b | 0 | IIIA | − |
| CC16018 | 81-85 | Male | Ascending | 42×30 | 3 | SS | X | 0 | IIA | − |
| CC16019 | 71-75 | Male | Transverse | 110×103 | 4a | SE | 0 | 0 | IIB | − |
| CC16020 | 71-75 | Female | Sigmoid | 35×70 | 3 | SS | 1a | 0 | IIIB | − |
| CC16021 | 71-75 | Male | Ascending | 39×42 | 3 | SS | 0 | 0 | IIA | − |
| CC16022 | 56-60 | Male | Rectum | 50×42 | 2 | MP | 0 | 0 | I | − |
| CC16023 | 81-85 | Female | Cecum | 55×82 | 3 | SS | 2b | 0 | IIIC | − |
| CC16024 | 36-40 | Female | Rectum | 61×45 | 3 | SS | 0 | 0 | IIA | − |
| CC16025 | 71-75 | Male | Ascending | 37×26 | 3 | SS | 0 | 0 | IIA | − |
| CC16026 | 76-80 | Female | Ascending | 58×49 | 3 | SS | 0 | 0 | IIA | − |
| CC16027 | 76-80 | Female | Ascending | 34×29 | 3 | SS | 1 | 0 | IIIA | − |
| CC16028 | 51-55 | Female | Sigmoid | 20×17 | 2 | MP | 0 | 0 | I | − |
| CC16029 | 56-60 | Female | Sigmoid | 46×42 | 3 | SS | 1a | 0 | IIIB | − |
| CC16030 | 66-70 | Male | Ascending | 17×20 | 3 | SS | 1b | 0 | IIIB | − |
| CC16031 | 76-80 | Male | Sigmoid | 33×30 | 1 | CIS | 0 | 0 | I | − |

| | | | | | | | | | | |
|---|---|---|---|---|---|---|---|---|---|---|
| CC16032 | 71-75 | Female | Ascending | 73×63 | 4a | SE | 1b | 0 | IIIB | − |
| CC16034 | 76-80 | Female | Sigmoid | 43×60 | 3 | SS | 1a | 0 | IIIB | − |
| CC16035 | 61-65 | Male | Rectum | 21×17 | 3 | SS | 2a | 0 | IIIB | − |
| CC16036 | 81-85 | Female | Cecum | 27×26 | 2 | 1b | 0 | 0 | IIIA | − |
| CC16037 | 51-55 | Female | Sigmoid | 58×32 | 3 | SS | 0 | 0 | IIA | − |
| CC16038 | 61-65 | Male | Ascending | 32×26 | 3 | SS | 0 | 0 | IIA | − |
| CC16039 | 81-85 | Male | Transverse | 27×26 | 3 | SS | 0 | 0 | IIA | − |
| CC16040 | 71-75 | Female | Sigmoid | 25×22 | 3 | SS | 0 | 0 | IIA | − |
| CC16041 | 66-70 | Male | Rectum | 35×42 | 3 | SS | 2a | 0 | IIIB | − |
| CC16042 | 66-70 | Female | Sigmoid | 54×33 | 3 | SS | 2a | 0 | IIIB | − |
| CC16043 | 56-60 | Female | Transverse | 47×28 | 3 | SS | 2a | 0 | IIIB | − |
| CC16044 | 71-75 | Male | Rectum | 52×35 | 3 | SS | 0 | 0 | IIA | − |
| CC16046 | 76-80 | Male | Sigmoid | 40×37 | 3 | SS | 0 | 0 | IIA | − |
| CC16047 | 66-70 | Female | Rectum | 57×32 | 3 | SS | 0 | 0 | IIA | − |
| CC16048 | 66-70 | Female | Sigmoid | 58×32 | 3 | SS | 0 | 0 | IIA | − |
| CC16049 | 81-85 | Female | Sigmoid | 53×52 | 3 | SS | 0 | 0 | IIA | − |
| CC16050 | 66-70 | Female | Transverse | 47×30 | 3 | SS | 0 | 0 | IIA | − |
| CC16051 | 66-70 | Female | Rectum | 88×62 | 3 | SS | 0 | 0 | IIA | − |
| CC16054 | 66-70 | Male | Ascending | 35×55 | 3 | SS | 1a | 0 | IIIB | − |
| CC16056 | 56-60 | Male | Rectum | 43×60 | 4b | SE | 2b | 0 | IIIC | − |
| CC16057 | 61-65 | Male | Transverse | 35×52 | 4b | SE | 2a | 0 | IIIC | − |
| CC16058 | 66-70 | Male | Rectum | 63×48 | 3 | SS | 0 | 0 | IIA | − |
| CC16060 | 61-65 | Male | Rectum | 80×100 | 3 | SS | 0 | 0 | IIA | − |

[a] TNM classification, 8th edition

[b] Pathological stage

[c] Nonregional abdominal para-aortic lymph nodes

**Supplementary Table S2. Primary tumor mutations detected through sequence analysis of Set 2 samples using Ion Proton™ and circulating tumor DNA levels (ctDNA) measured by digital PCR.**

| Case ID | Chromosomal location | Gene | Variant type | Base change | Amino acid change | Total allele | Wild type | Mutant allele | Primary VAF[a] (%) | dPCR[b] | pre ctDNA[c] (%) |
|---|---|---|---|---|---|---|---|---|---|---|---|
| CC16002 | chr12:25398280 | *KRAS* | missense | c.37G>T | p.G13C | 1995 | 1790 | 205 | 10.3 | yes | 0 |
| CC16003 | chrX:63411678 | *AMER1* | nonsense | c.1489C>T | p.R497* | 1997 | 1385 | 612 | 30.6 | | |
| | chr5:112175639 | *APC* | nonsense | c.4348C>T | p.R1450* | 1999 | 1021 | 978 | 48.9 | | |
| | chr17:7578256 | *TP53* | nonsense | c.586C>T | p.R196* | 1948 | 1025 | 923 | 47.4 | yes | 0.43 |
| CC16006 | chr14:21868397 | *CHD8* | missense | c.4640A>G | p.H1547R | 1998 | 1586 | 412 | 20.62 | | |
| | chr12:25398283 | *KRAS* | missense | c.34G>T | p.G12C | 1998 | 1779 | 218 | 10.91 | yes | 0 |
| | chr17:7578368 | *TP53* | splicesite_3 | c.559+3G>C | | 1920 | 1543 | 376 | 19.6 | | |
| CC16009 | chr17:63554007 | *AXIN2* | frameshift deletion | c.731delC | p.S244fs | 1991 | 1228 | 763 | 38.3 | | |
| | chr12:25398283 | *KRAS* | missense | c.35G>A | p.G12D | 1997 | 1459 | 538 | 26.9 | yes | 0.25 |
| | chr2:48027519 | *MSH6* | missense | c.2397G>A | p.M799I | 2000 | 1677 | 323 | 16.2 | | |
| | chr17:7578368 | *TP53* | splicesite_3 | c.559+3G>C | | 1910 | 1274 | 635 | 33.2 | | |
| CC16017 | chr5:112175639 | APC | nonsense | c.4348C>T | p.R1450* | 1996 | 1081 | 915 | 45.8 | | |
| | chr5:112173914 | APC | nonsense | c.2623A>T | p.K875* | 1999 | 1167 | 832 | 41.6 | | |
| | chr3:41278106 | CTNNB1 | missense | c.1982G>T | p.R661L | 1998 | 1101 | 897 | 44.9 | | |
| | chr12:25378561 | KRAS | missense | c.437C>T | p.A146V | 1550 | 869 | 681 | 43.9 | | |

| | | | | | | | | | | | |
|---|---|---|---|---|---|---|---|---|---|---|---|
| | chr1:115258745 | NRAS | missense | c.37G>C | p.G13R | 2000 | 1492 | 506 | 25.3 | | |
| | chr3:178936082 | PIK3CA | missense | c.1624G>A | p.E542K | 1318 | 1017 | 301 | 22.8 | yes | 0 |
| | chr5:112175672 | APC | frameshift deletion | c.4391_4394delAGAG | p.E1464fs | 1949 | 1525 | 423 | 21.70 | | |
| CC16018 | chr5:112174631 | APC | nonsense | c.3340C>T | p.R1114* | 1999 | 1589 | 410 | 20.51 | | |
| | chr17:7577546 | TP53 | missense | c.733G>A | p.G245S | 1993 | 1421 | 570 | 28.60 | yes | 0.18 |
| CC16019 | chr7:140453133 | BRAF | missense | c.1799T>A | p.V600E | 1980 | 1632 | 347 | 17.53 | yes | 11.9 |
| | chr17:7577120 | TP53 | missense | c.818G>A | p.R273H | 1966 | 1165 | 801 | 40.74 | yes | 4.57 |
| CC16020 | chr5:112151261 | APC | missense | c.904C>T | p.R302* | 1998 | 1107 | 891 | 44.59 | yes | 0.17 |
| | chr17:7578474 | TP53 | missense | c.451C>G | p.P151A | 1616 | 809 | 803 | 49.69 | yes | 0.75 |
| | chr7:140453155 | BRAF | missense | c.1780G>C | p.D594H | 1991 | 1507 | 484 | 24.31 | yes | 0 |
| CC16021 | chr20:57485014 | GNAS | missense | c.848G>A | p.R283H | 1998 | 1556 | 442 | 22.12 | | |
| | chr12:25378562 | KRAS | missense | c.436G>A | p.A146T | 1999 | 1762 | 237 | 11.86 | | |
| | chr5:112128203 | APC | nonsense | c.706C>T | p.Q236* | 1999 | 1612 | 387 | 19.36 | | |
| | chr4:153247376 | FBXW7 | nonframeshift deletion | c.1423_1425delGTT | p.V475del | 1984 | 1114 | 870 | 43.85 | | |
| CC16022 | chr4:153332760 | FBXW7 | frameshift insertion | c.195_196insGA | p.P66fs | 1997 | 1522 | 475 | 23.79 | | |
| | chr12:25398280 | KRAS | missense | c.38G>A | p.G13D | 1997 | 1050 | 944 | 47.27 | yes | 0.55 |
| | chr17:7578474 | TP53 | missense | c.451C>A | p.P151T | 1978 | 782 | 1194 | 60.36 | | |
| CC16024 | chr2:148683611 | ACVR2A | nonsense | c.1228G>T | p.E410* | 1893 | 1278 | 615 | 32.49 | | |

| | | | | | | | | | | |
|---|---|---|---|---|---|---|---|---|---|---|
| chr5:112175046 | APC | missense | c.3755C>A | p.S1252Y | 1914 | 862 | 653 | 34.12 | | |
| chr1:27106178 | ARID1A | nonsense | c.5789C>A | p.S1930* | 1092 | 807 | 285 | 26.10 | | |
| chr11:108202715 | ATM | missense | c.7739G>T | p.R2580I | 1997 | 1418 | 579 | 28.99 | | |
| chr11:108196939 | ATM | missense | c.6962C>T | p.A2321V | 1998 | 1471 | 527 | 26.38 | | |
| chr14:21871721 | CHD8 | missense | c.3409C>A | p.L1137I | 1999 | 1461 | 538 | 26.91 | | |
| chr14:21860965 | CHD8 | missense | c.6472C>T | p.R2158C | 1993 | 1491 | 502 | 25.19 | | |
| chr3:41277291 | CTNNB1 | missense | c.1760G>A | p.R587Q | 1998 | 1352 | 646 | 32.33 | | |
| chr3:41266232 | CTNNB1 | nonsense | c.229G>T | p.E77* | 1994 | 1441 | 553 | 27.73 | | |
| chr8:42231798 | DKK4 | missense | c.495T>G | p.I165M | 1431 | 1172 | 259 | 18.10 | | |
| chr4:153244184 | FBXW7 | missense | c.1973G>A | p.R658Q | 1977 | 1405 | 572 | 28.93 | | |
| chr4:153332915 | FBXW7 | missense | c.41G>A | p.R14Q | 1979 | 1463 | 516 | 26.07 | | |
| chr12:130648241 | FZD10 | missense | c.754G>A | p.D252N | 1991 | 1407 | 584 | 29.33 | | |
| chr12:130648713 | FZD10 | missense | c.1226G>A | p.G409D | 1999 | 1457 | 542 | 27.11 | | |
| chr12:130647576 | FZD10 | missense | c.89G>A | p.G30D | 1844 | 1509 | 335 | 18.17 | | |
| chr3:119624687 | GSK3B | missense | c.728C>T | p.A243V | 694 | 507 | 187 | 26.95 | | |
| chr2:48027661 | MSH6 | nonsense | c.2539G>T | p.E847* | 203 | 129 | 74 | 36.45 | yes | 0 |
| chr3:178916716 | PIK3CA | nonsense | c.103G>T | p.E35* | 1391 | 972 | 419 | 30.12 | | |
| chr3:178922301 | PIK3CA | missense | c.1070G>A | p.R357Q | 2000 | 1446 | 554 | 27.70 | | |
| chr3:178916642 | PIK3CA | missense | c.29T>G | p.L10R | 2000 | 1593 | 407 | 20.35 | | |

| | | | | | | | | | | | |
|---|---|---|---|---|---|---|---|---|---|---|---|
| | chr10:89692895 | PTEN | missense | c.389G>A | p.R130Q | 1972 | 1462 | 510 | 25.86 | | |
| | chr17:56435626 | RNF43 | missense | c.1511A>G | p.D504G | 1999 | 1729 | 270 | 13.51 | | |
| | chr18:48603033 | SMAD4 | missense | c.1334G>A | p.R445Q | 1402 | 1023 | 379 | 27.03 | | |
| | chr17:6676434 | XAF1 | missense | c.852G>T | p.E284D | 365 | 304 | 61 | 16.71 | | |
| CC16025 | chrX:63410048 | AMER1 | missense | c.3119C>A | p.A1040D | 2000 | 1773 | 227 | 11.35 | yes | 0 |
| | chr20:57484420 | *GNAS* | missense | c.601C>T | p.R201C | 1999 | 1441 | 558 | 27.91 | | |
| | chr12:25378562 | *KRAS* | missense | c.436G>A | p.A146T | 1999 | 590 | 1409 | 70.49 | | |
| CC16028 | chr3:178936091 | *PIK3CA* | missense | c.1633G>A | p.E545K | 1999 | 1161 | 838 | 41.92 | yes | 0 |
| | chr10:114912175 | *TCF7L2* | frameshift deletion | c.1247delG | p.G416fs | 1991 | 946 | 1045 | 52.49 | | |
| | chr17:7578443 | *TP53* | missense | c.476C>T | p.A159V | 1932 | 596 | 1336 | 69.15 | | |
| | chr5:112173917 | *APC* | nonsense | c.2626C>T | p.R876* | 2000 | 1535 | 465 | 23.25 | yes | 0 |
| CC16029 | chr5:112175628 | *APC* | frameshift deletion | c.4338delT | p.Q1447fs | 1987 | 1533 | 454 | 22.85 | | |
| | chr4:153258983 | *FBXW7* | nonsense | c.832C>T | p.R278* | 1653 | 1285 | 368 | 22.26 | | |
| | chr17:7578490 | *TP53* | nonsense | c.438G>A | p.W146* | 1824 | 1203 | 621 | 34.05 | | |
| | chrX:63411969 | *AMER1* | nonsense | c.1198G>T | p.E400* | 1568 | 484 | 1084 | 69.13 | | |
| | chr5:112176020 | *APC* | nonsense | c.4729G>T | p.E1577* | 959 | 633 | 326 | 33.99 | | |
| CC16030 | chr5:112173736 | *APC* | frameshift Insertion | c.2454_2455insTGGCAAC | p.M819fs | 1996 | 1641 | 355 | 17.79 | | |
| | chr12:25378561 | *KRAS* | missense | c.437C>T | p.A146V | 1993 | 728 | 1265 | 63.47 | yes | 0.51 |
| | chr10:89720665 | *PTEN* | nonsense | c.821G>A | p.W274* | 212 | 122 | 90 | 42.45 | | |

| | | | | | | | | | | | |
|---|---|---|---|---|---|---|---|---|---|---|---|
| | chr17:7577021 | *TP53* | nonsense | c.916C>T | p.R306* | 2000 | 1100 | 900 | 45.00 | yes | 0 |
| CC16031 | chr17:70117936 | *SOX9* | missense | c.404T>A | p.L135H | 1997 | 1762 | 235 | 11.77 | yes | 0.06 |
| CC16036 | chrX:63412020 | *AMER1* | nonsense | c.1147G>T | p.E383* | 1999 | 1385 | 614 | 30.72 | yes | 0.22 |
| | chr5:112174682 | *APC* | nonsense | c.3391C>T | p.Q1131* | 1998 | 1629 | 369 | 18.47 | | |
| | chr12:25398280 | *KRAS* | missense | c.38G>A | p.G13D | 1996 | 1534 | 462 | 23.15 | | |
| | chr11:68179025 | *LRP5* | missense | c.2440G>A | p.A814T | 1994 | 1754 | 240 | 12.04 | | |
| | chr3:178936094 | *PIK3CA* | missense | c.1636C>A | p.Q546K | 545 | 408 | 137 | 25.14 | | |
| | chr17:7577156 | *TP53* | splicesite_5 | c.783-1G>A | | 1972 | 1348 | 624 | 31.64 | | |
| CC16037 | chr5:112170648 | *APC* | nonsense | c.1744G>T | p.E582* | 2000 | 1680 | 320 | 16.00 | | |
| | chr17:7577556 | *TP53* | missense | c.722C>G | p.S241C | 1978 | 1187 | 788 | 39.84 | yes | 0.27 |
| CC16038 | chr12:25398280 | *KRAS* | missense | c.38G>A | p.G13D | 1998 | 1822 | 176 | 8.81 | yes | 0 |
| CC16039 | chr15:65116026 | *PIF1* | missense | c.509C>T | p.T170M | 813 | 656 | 157 | 19.31 | | |
| | chr17:7577553 | *TP53* | frameshift Insertion | c.714_715insGT | p.N239fs | 1492 | 666 | 825 | 55.29 | yes | 0 |
| CC16040 | chr5:112173917 | *APC* | nonsense | c.2626C>T | p.R876* | 1997 | 857 | 1140 | 57.09 | yes | 0 |
| | chr1:27107138 | *ARID1A* | frameshift deletion | c.6750delG | p.E2250fs | 1998 | 1231 | 767 | 38.39 | | |
| | chr12:25398283 | *KRAS* | missense | c.35G>A | p.G12D | 1999 | 1255 | 744 | 37.22 | yes | 13.2 |
| | chr3:178936095 | *PIK3CA* | missense | c.1637A>G | p.Q546R | 697 | 273 | 424 | 60.83 | | |
| | chr17:7578256 | *TP53* | missense | c.584T>C | p.I195T | 1949 | 797 | 1152 | 59.11 | | |

| | | | | | | | | | | | |
|---|---|---|---|---|---|---|---|---|---|---|---|
| CC16044 | chr5:112116592 | *APC* | nonsense | c.637C>T | p.R213* | 1687 | 727 | 960 | 56.91 | yes | 1.73 |
| | chr4:153249511 | *FBXW7* | missense | c.1267G>A | p.G423R | 1998 | 1280 | 718 | 35.94 | | |
| CC16046 | chr5:112173718 | *APC* | frameshift deletion | c.2428delA | p.R810fs | 1995 | 1195 | 800 | 40.10 | | |
| | chr5:112128141 | *APC* | splicesite_5 | c.646-2A>G | | 1065 | 668 | 397 | 37.28 | | |
| | chr12:25398283 | *KRAS* | missense | c.35G>A | p.G12D | 1997 | 1209 | 784 | 39.26 | yes | 0 |
| | chr3:178952085 | *PIK3CA* | missense | c.3140A>G | p.H1047R | 1999 | 1210 | 788 | 39.42 | yes | 0.84 |
| CC16047 | chr5:112128143 | *APC* | nonsense | c.646C>T | p.R216* | 1083 | 393 | 690 | 63.71 | | |
| | chr15:65112165 | *PIF1* | missense | c.1214G>A | p.R405H | 1905 | 1357 | 548 | 28.77 | | |
| | chr3:178936091 | *PIK3CA* | missense | c.1633G>A | p.E545K | 671 | 452 | 219 | 32.64 | yes | 0 |
| CC16048 | chr7:140453133 | *BRAF* | missense | c.1799T>A | p.V600E | 1979 | 881 | 1096 | 55.38 | yes | 0 |
| | chr4:153258983 | *FBXW7* | nonsense | c.832C>T | p.R278* | 1999 | 1243 | 756 | 37.82 | | |
| | chr12:130648458 | *FZD10* | missense | c.971C>T | p.S324L | 2000 | 1229 | 771 | 38.55 | | |
| | chr17:7579348 | *TP53* | frameshift deletion | c.338delT | p.F113fs | 1992 | 1266 | 724 | 36.35 | | |
| CC16049 | chr12:25398283 | *KRAS* | missense | c.35G>T | p.G12V | 2000 | 1250 | 747 | 37.35 | yes | 0 |
| | chr17:7577528 | *TP53* | missense | c.742C>T | p.R248W | 1963 | 576 | 1386 | 70.61 | | |
| CC16050 | chr5:112174580 | *APC* | nonsense | c.3289G>T | p.E1097* | 1997 | 1214 | 783 | 39.21 | | |
| | chr14:21876575 | *CHD8* | nonsense | c.2626C>T | p.R876* | 1997 | 882 | 1115 | 55.83 | | |
| | chr4:153247367 | *FBXW7* | nonsense | c.1435C>T | p.R479* | 1765 | 1210 | 555 | 31.44 | | |

| | | | | | | | | | | |
|---|---|---|---|---|---|---|---|---|---|---|
| chr17:70117938 | *SOX9* | missense | c.406A>G | p.S136G | 2000 | 738 | 1262 | 63.10 | | |
| chr17:7577580 | *TP53* | missense | c.701A>G | p.Y234C | 1980 | 534 | 1446 | 73.03 | yes | 0 |

[a] Variant allele frequency

[b] Plasma DNA was evaluated by dPCR

[c] Pretreatment circulating tumor DNA

**Supplementary Table S3. Primary tumor mutations detected through sequence analysis of Set 3 samples using Ion 5S™ and circulating tumor DNA (ctDNA) levels measured by digital PCR.**

| Case ID | Chromosomal location | Gene | Variant type | Base change | Amino acid change | Total allele | Wild type | Mutant allele | Primary VAF[a] (%) | dPCR[b] | pre ctDNA[c] (%) |
|---|---|---|---|---|---|---|---|---|---|---|---|
| CC16010 | chr17:7675108 | *TP53* | frameshift insertion | c.107_108insAGCA | p.H36Qfs*14 | 4610 | 2983 | 981 | 21.11 | yes | 8.54 |
| CC16012 | chr5:112839729 | *APC* | stopgain | c.G4135T | p.E1379* | 3959 | 2075 | 1880 | 47.49 | | |
| | chr12:25245350 | *KRAS* | nonsynonymous | c.G35T | p.G12V | 7680 | 3872 | 3802 | 49.51 | yes | 0.51 |
| | chr20:37035354 | *RBL1* | frameshift insertion | c.776dupC | p.S260Ffs*6 | 4520 | 3858 | 1198 | 23.58 | | |
| | chr17:7675232 | *TP53* | nonsynonymous | c.C380A | p.S127Y | 3021 | 605 | 2384 | 78.91 | yes | 0.19 |
| CC16013 | chr11:108335029 | *ATM* | nonsynonymous | c.C8071T | p.R2691C | 3427 | 2342 | 983 | 29.53 | | |
| | chr7:140753336 | *BRAF* | nonsynonymous | c.T1799A | p.V600E | 5149 | 3438 | 1697 | 32.95 | yes | 0 |
| | chr20:58855061 | *GNAS* | nonsynonymous | c.G1796A | p.R599H | 4196 | 2611 | 1576 | 37.55 | | |
| | chr3:179234297 | *PIK3CA* | nonsynonymous | c.A3140G | p.H1047R | 5276 | 3295 | 1969 | 37.26 | yes | 0 |
| | chr17:58362564 | *RNF43* | nonsynonymous | c.C667T | p.R223C | 2587 | 1341 | 1245 | 48.13 | | |
| | chr10:113165642 | *TCF7L2* | frameshift deletion | c.1411delC | p.S473Pfs*5 | 2148 | 1442 | 671 | 30.74 | | |
| CC16014 | chr5:112839693 | *APC* | frameshift insertion | c.4046dupA | p.T1350Dfs*7 | 2540 | 1707 | 837 | 32.66 | yes | 8.3 |
| | chr14:21393628 | *CHD8* | nonsynonymous | c.G6167A | p.R2056Q | 2700 | 1565 | 1112 | 41.12 | | |
| | chr17:7675131 | *TP53* | nonsynonymous | c.G481A | p.A161T | 2370 | 1545 | 814 | 34.39 | yes | 8.68 |
| CC16015 | chr11:108229288 | *ATM* | nonsynonymous | c.G296A | p.S99N | 2630 | 2047 | 558 | 21.17 | | |

| | | | | | | | | | | | |
|---|---|---|---|---|---|---|---|---|---|---|---|
| | chr11:108229289 | *ATM* | nonsynonymous | c.T297G | p.S99R | 2543 | 1999 | 543 | 21.36 | | |
| | chr7:140800366 | *BRAF* | nonsynonymous | c.A976G | p.I326V | 6118 | 2861 | 3254 | 53.17 | | |
| | chr3:37025979 | *MLH1* | stopgain | c.A658T | p.K220* | 2536 | 1912 | 620 | 24.45 | | |
| | chr17:58358121 | *RNF43* | nonsynonymous | c.G1274A | p.R425H | 7705 | 5986 | 1689 | 21.95 | | |
| | chr17:58358788 | *RNF43* | stopgain | c.C988T | p.R330* | 1226 | 249 | 32 | 11.27 | yes | 2.43 |
| | chr17:58363351 | *RNF43* | nonsynonymous | c.C125A | p.A42D | 2536 | 1912 | 620 | 24.45 | | |
| CC16026 | chr5:112775687 | *APC* | stopgain | c.C481T | p.Q161* | 2430 | 1392 | 1027 | 42.21 | | |
| | chr12:25245350 | *KRAS* | nonsynonymous | c.G35A | p.G12D | 7144 | 5172 | 1967 | 27.54 | yes | 0 |
| CC16027 | chr5:112839942 | *APC* | stopgain | c.C4348T | p.R1450* | 3185 | 2036 | 1148 | 36.02 | | |
| | chr12:25245350 | *KRAS* | nonsynonymous | c.G35T | p.G12V | 5261 | 3070 | 2188 | 41.6 | yes | 0 |
| | chr17:7675180 | *TP53* | frameshift insertion | c.35_36insTGCA | p.Q12Hfs*6 | 2962 | 1151 | 1804 | 60.42 | yes | 0.27 |
| CC16032 | chr5:112839879 | *APC* | stopgain | c.C4285T | p.Q1429* | 2611 | 1315 | 1265 | 48.37 | | |
| | chr1:26772550 | *ARID1A* | frameshift deletion | c.3458delC | p.M1154Wfs*7 | 2596 | 1311 | 1291 | 49.43 | | |
| | chr17:65537787 | *AXIN2* | nonsynonymous | c.G1249T | p.A417S | 2424 | 1763 | 518 | 22.7 | | |
| | chr1:114713909 | *NRAS* | nonsynonymous | c.C181A | p.Q61K | 3255 | 2100 | 1053 | 33.31 | yes | 21.6 |
| | chr3:179218303 | *PIK3CA* | nonsynonymous | c.G1633A | p.E545K | 1015 | 766 | 249 | 24.53 | yes | 11.8 |
| | chr18:51078425 | *SMAD4* | frameshift deletion | c.1618delC | p.L540Ffs*12 | 2693 | 1435 | 1279 | 47.02 | | |
| | chr17:72121597 | *SOX9* | frameshift deletion | c.207delC | p.V71Cfs*39 | 2040 | 1510 | 462 | 22.42 | | |
| | chr17:7673763 | *TP53* | frameshift deletion | c.460delG | p.E154Kfs*59 | 1751 | 856 | 899 | 51.17 | yes | 0.19 |

| | | | | | | | | | | | |
|---|---|---|---|---|---|---|---|---|---|---|---|
| CC16051 | chr5:112839117 | *APC* | stopgain | c.C3523T | p.Q1175* | 4098 | 3380 | 707 | 17.21 | | |
| | chr17:7675088 | *TP53* | nonsynonymous | c.G524A | p.R175H | 7057 | 6177 | 872 | 12.34 | yes | 0.31 |
| CC16054 | chrX:64192215 | *AMER1* | stopgain | c.C1072T | p.R358* | 3751 | 770 | 2980 | 79.42 | | |
| | chrX:64191813 | *AMER1* | frameshift insertion | c.1473dupC | p.R492Qfs*16 | 1573 | 1447 | 372 | 20.26 | | |
| | chrX:64191986 | *AMER1* | frameshift insertion | c.1300dupC | p.H434Pfs*7 | 499 | 265 | 268 | 49.91 | | |
| | chrX:64192785 | *AMER1* | frameshift insertion | c.501dupA | p.G168Rfs*7 | 287 | 141 | 193 | 57.78 | | |
| | chrX:64193123 | *AMER1* | frameshift insertion | c.163dupA | p.T55Nfs*17 | 2999 | 2365 | 707 | 22.91 | | |
| | chr5:112838455 | *APC* | stopgain | c.T2861A | p.L954* | 4502 | 2996 | 1494 | 33.11 | | |
| | chr5:112840155 | *APC* | stopgain | c.G4561T | p.E1521* | 3346 | 2651 | 682 | 20.36 | | |
| | chr5:112840372 | *APC* | nonsynonymous | c.A4724G | p.K1575R | 645 | 566 | 77 | 11.94 | | |
| | chr5:112841875 | *APC* | nonsynonymous | c.C6281T | p.P2094L | 4071 | 3606 | 461 | 11.32 | | |
| | chr5:112819329 | *APC* | frameshift insertion | c.1244dupA | p.D416Gfs*10 | 941 | 711 | 296 | 29.37 | | |
| | chr5:112840383 | *APC* | frameshift insertion | c.4736dupC | p.A1580Cfs*34 | 588 | 484 | 191 | 28.25 | | |
| | chr1:26773683 | *ARID1A* | nonsynonymous | c.T3970C | p.Y1324H | 1768 | 1573 | 187 | 10.56 | | |
| | chr1:26773685 | *ARID1A* | stopgain | c.C3972A | p.Y1324* | 1482 | 1238 | 204 | 14.14 | | |
| | chr1:26779587 | *ARID1A* | nonsynonymous | c.C5689G | p.P1897A | 1764 | 903 | 370 | 28.95 | | |
| | chr1:26774569 | *ARID1A* | frameshift insertion | c.4343dupC | p.G1449Rfs*42 | 546 | 297 | 297 | 49.25 | | |
| | chr1:26774597 | *ARID1A* | frameshift insertion | c.4371dupC | p.Q1458Pfs*33 | 634 | 390 | 267 | 40.39 | | |
| | chr1:26774620 | *ARID1A* | frameshift insertion | c.4394dupC | p.A1466Cfs*25 | 830 | 643 | 236 | 26.85 | | |

| | | | | | | | | | | |
|---|---|---|---|---|---|---|---|---|---|---|
| chr11:108244102 | *ATM* | frameshift insertion | c.646_647insT | p.A216Vfs*38 | 1826 | 1183 | 626 | 33.23 | | |
| chr11:108281146 | *ATM* | frameshift insertion | c.3555dupA | p.E1186Rfs*14 | 850 | 547 | 349 | 38.86 | | |
| chr8:60852937 | *CHD7* | frameshift deletion | c.6213delC | p.Q2073Sfs*71 | 1740 | 1153 | 561 | 31.22 | | |
| chr8:60865588 | *CHD7* | frameshift insertion | c.2503dupC | p.S836Lfs*56 | 1880 | 801 | 1108 | 57.29 | | |
| chr14:21391024 | *CHD8* | stopgain | c.A7105T | p.K2369X | 1101 | 937 | 150 | 13.57 | | |
| chr20:58853838 | *GNAS* | nonsynonymous | c.T386C | p.L129P | 2997 | 1550 | 1441 | 48.03 | | |
| chr12:25245350 | *KRAS* | nonsynonymous | c.G35A | p.G12D | 7392 | 3692 | 3700 | 50.05 | yes | 0 |
| chr12:25225691 | *KRAS* | frameshift insertion | c.372dupA | p.V125Sfs*19 | 837 | 601 | 337 | 35.93 | | |
| chr11:68390006 | *LRP5* | frameshift insertion | c.1539dupG | p.E514Gfs*20 | 818 | 603 | 244 | 28.74 | | |
| chr11:68403698 | *LRP5* | frameshift insertion | c.1801dupG | p.T602Nfs*35 | 317 | 235 | 88 | 27.16 | | |
| chr11:68425992 | *LRP5* | frameshift insertion | c.1699_1700insG | p.T567Sfs*73 | 1545 | 953 | 714 | 41.34 | | |
| chr3:37025946 | *MLH1* | nonsynonymous | c.G625A | p.D209N | 2946 | 2222 | 721 | 24.47 | | |
| chr3:37025947 | *MLH1* | nonsynonymous | c.A626G | p.D209G | 4189 | 3242 | 725 | 17.1 | | |
| chr3:37050487 | *MLH1* | stopgain | c.1899dupT | p.E634* | 2225 | 1589 | 1217 | 43.28 | | |
| chr3:179204536 | *PIK3CA* | nonsynonymous | c.G1093A | p.E365K | 1706 | 1190 | 508 | 29.85 | | |
| chr10:87952121 | *PTEN* | frameshift insertion | c.497dupT | p.T167Nfs*13 | 3258 | 2607 | 854 | 24.64 | | |
| chr17:72124335 | *SOX9* | frameshift insertion | c.1479dupC | p.S494Qfs*84 | 3009 | 2262 | 857 | 27.25 | | |
| chr10 :13141234 | *TCF7L2* | frameshift insertion | c.175dupC | p.L60Sfs*7 | 2834 | 2365 | 1314 | 35.39 | | |
| chr17:7675140 | *TP53* | nonsynonymous | c.C472T | p.R158C | 2243 | 1395 | 846 | 37.72 | | |

| | | | | | | | | | | | |
|---|---|---|---|---|---|---|---|---|---|---|---|
| CC16056 | chr5:112838007 | *APC* | stopgain | c.C2413T | p.R805* | 3842 | 3363 | 473 | 12.32 | | |
| | chr12:25245350 | *KRAS* | nonsynonymous | c.G35C | p.G12A | 7850 | 5479 | 2367 | 30.15 | yes | 0.43 |
| | chr17:7673776 | *TP53* | nonsynonymous | c.C844T | p.R282W | 2675 | 1866 | 807 | 30.17 | yes | 0.11 |
| CC16057 | chr17:7674894 | *TP53* | stopgain | c.C637T | p.R213* | 6914 | 5266 | 1456 | 21.14 | yes | 0 |
| | chr17:7675237 | *TP53* | splicing | c.376-1G>A | | 2716 | 2013 | 703 | 25.87 | | |
| CC16058 | chr2:147926138 | *ACVR2A* | nonsynonymous | c.A1000G | p.R334G | 828 | 721 | 107 | 12.92 | | |
| | chrX:64192583 | *AMER1* | nonsynonymous | c.C704T | p.P235L | 250 | 42 | 34 | 44.74 | | |
| | chr5:112780895 | *APC* | stopgain | c.C637T | p.R213* | 3101 | 2512 | 583 | 18.78 | | |
| | chr7:140924640 | *BRAF* | nonsynonymous | c.G64A | p.D22N | 3979 | 1662 | 2299 | 58.04 | | |
| | chr4:152328232 | *FBXW7* | nonsynonymous | c.G1394A | p.R465H | 7171 | 5373 | 1796 | 25.03 | | |
| | chr12:25245350 | *KRAS* | nonsynonymous | c.G35C | p.G12A | 7807 | 6214 | 1591 | 20.38 | yes | 0 |
| | chr17:7673775 | *TP53* | nonframeshift insertion | c.448_449insACC | p.D149_R150insH | 3013 | 2195 | 813 | 26.89 | yes | 2.05 |
| CC16060 | chr17:7673802 | *TP53* | nonsynonymous | c.G818A | p.R273H | 6995 | 1890 | 5098 | 72.85 | yes | 1.22 |

[a] Variant allele frequency,

[b] Plasma DNA was evaluated by dPCR,

[c] Variant circulating tumor DNA

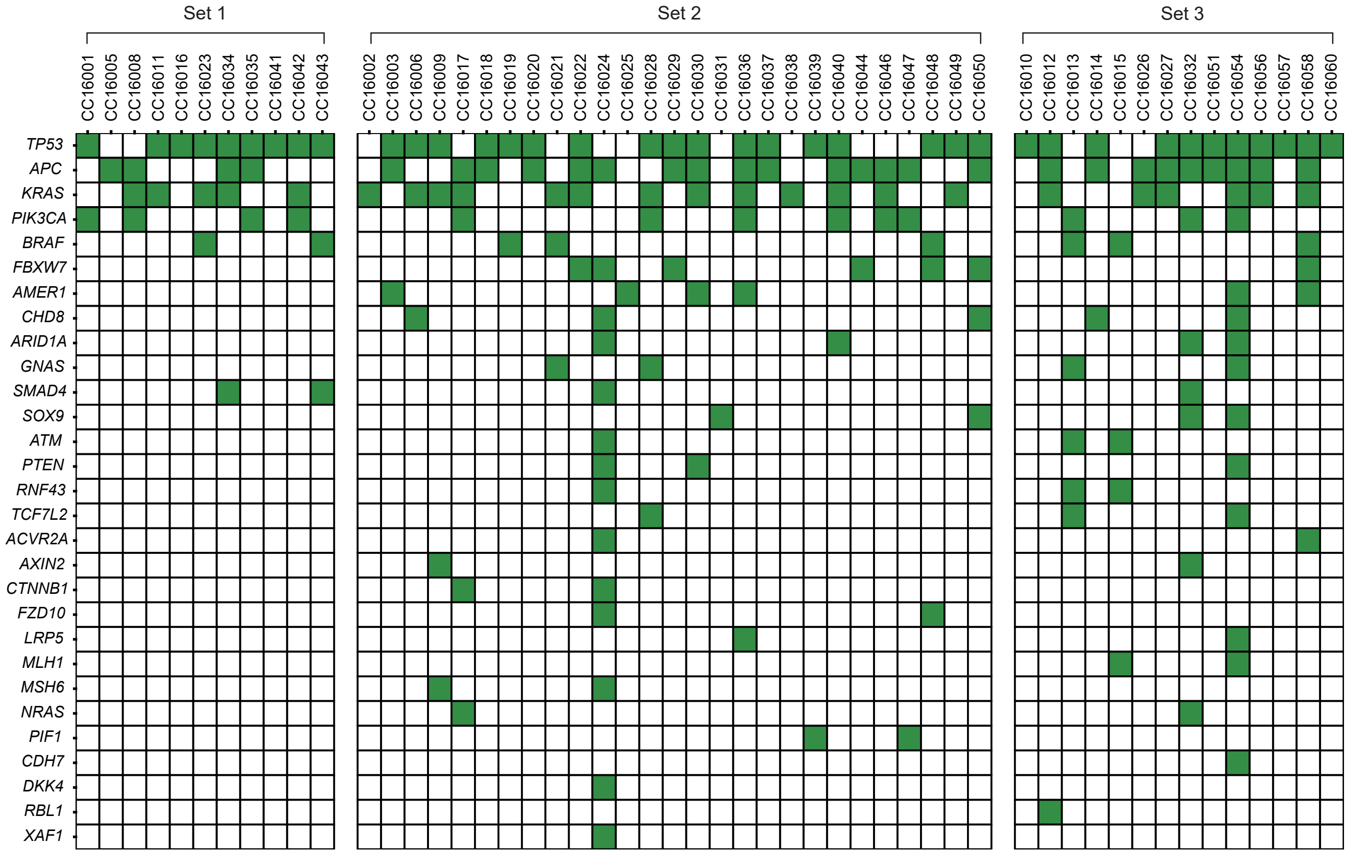

**Supplementary Figure S1. Somatic mutation profile of primary CRC tumors from the 52 patients in the current study cohort.**

Mutated genes that satisfied the criteria of each set are shown. The top row indicates the patient ID. Genes listed in the custom panel are arranged on the left side of the panel. The detailed mutation profile for Set 1 is available in our previous report (Ref. 35 in the main text). Green boxes indicate the presence of mutations.

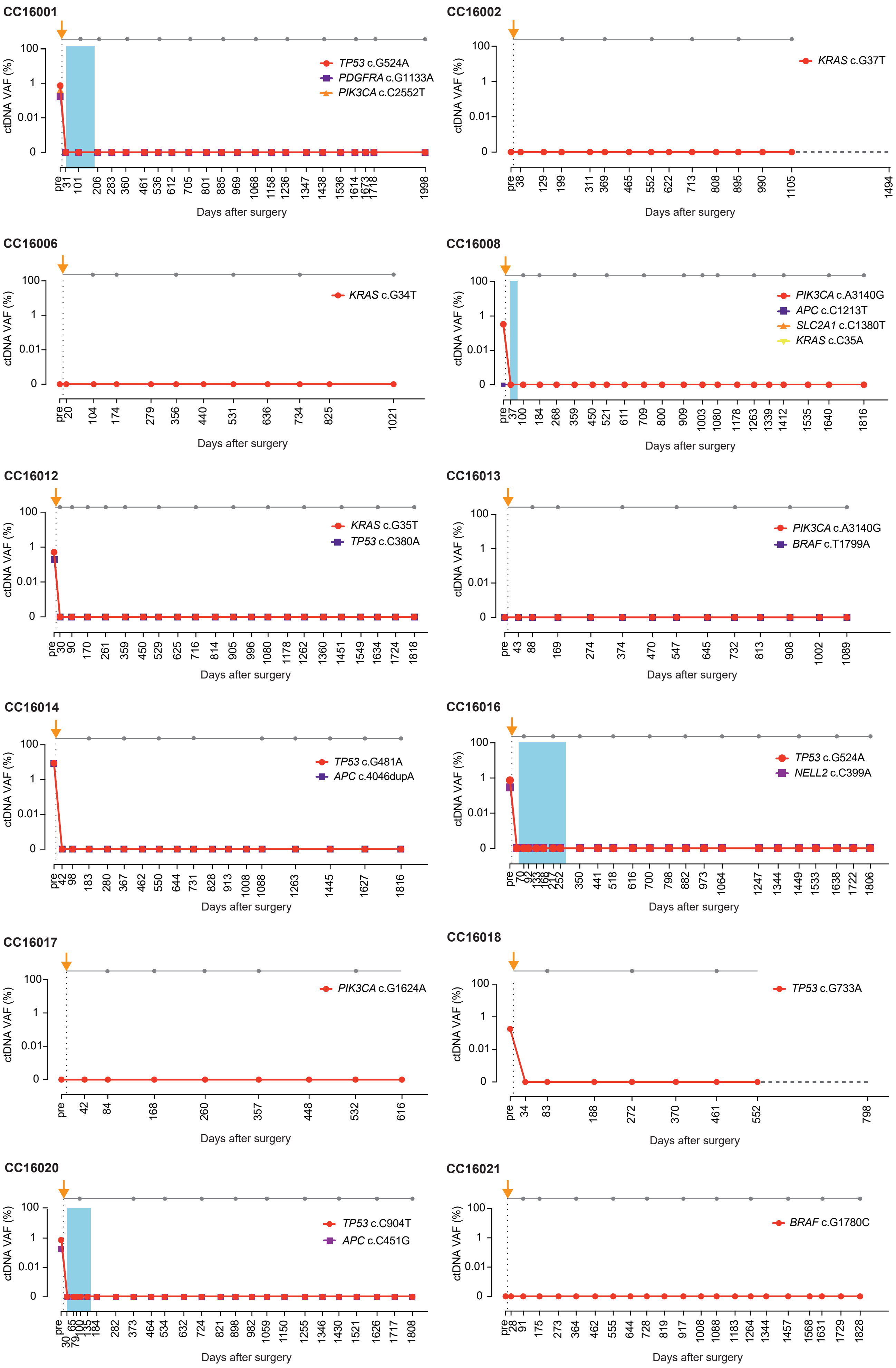
CC16001
TP53 c.G524A
PDGFRA c.G1133A
PIK3CA c.C2552T
CC16002
KRAS c.G37T
CC16006
KRAS c.G34T
CC16008
PIK3CA c.A3140G
APC c.C1213T
SLC2A1 c.C1380T
KRAS c.C35A
CC16012
KRAS c.G35T
TP53 c.C380A
CC16013
PIK3CA c.A3140G
BRAF c.T1799A
CC16014
TP53 c.G481A
APC c.4046dupA
CC16016
TP53 c.G524A
NELL2 c.C399A
CC16017
PIK3CA c.G1624A
CC16018
TP53 c.G733A
CC16020
TP53 c.C904T
APC c.C451G
CC16021
BRAF c.G1780C
ctDNA VAF (%)
100
1
0.01
0
Days after surgery

continues

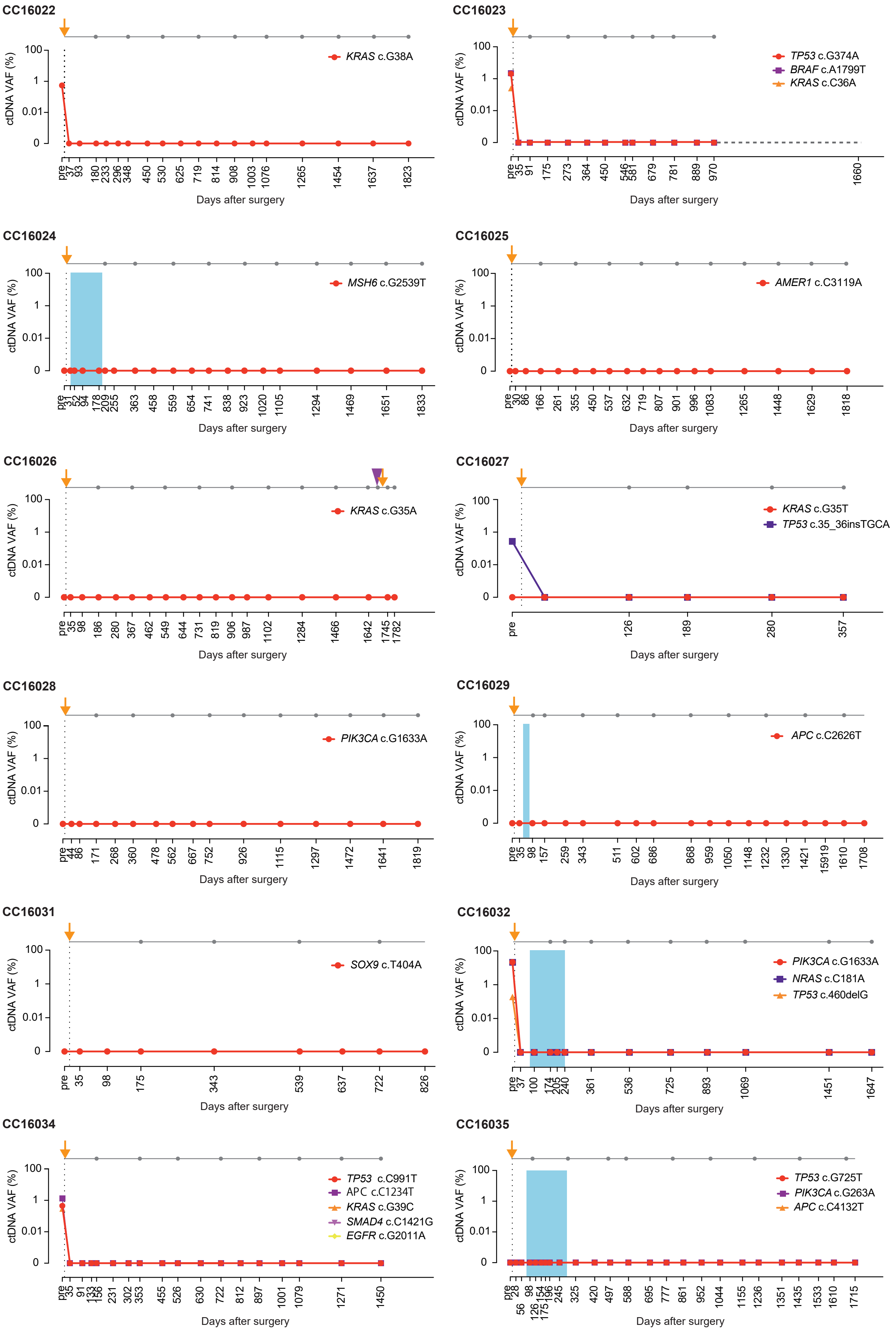
CC16022
ctDNA VAF (%)
KRAS c.G38A
Days after surgery
CC16023
ctDNA VAF (%)
TP53 c.G374A
BRAF c.A1799T
KRAS c.C36A
Days after surgery
CC16024
ctDNA VAF (%)
MSH6 c.G2539T
Days after surgery
CC16025
ctDNA VAF (%)
AMER1 c.C3119A
Days after surgery
CC16026
ctDNA VAF (%)
KRAS c.G35A
Days after surgery
CC16027
ctDNA VAF (%)
KRAS c.G35T
TP53 c.35_36insTGCA
Days after surgery
CC16028
ctDNA VAF (%)
PIK3CA c.G1633A
Days after surgery
CC16029
ctDNA VAF (%)
APC c.C2626T
Days after surgery
CC16031
ctDNA VAF (%)
SOX9 c.T404A
Days after surgery
CC16032
ctDNA VAF (%)
PIK3CA c.G1633A
NRAS c.C181A
TP53 c.460delG
Days after surgery
CC16034
ctDNA VAF (%)
TP53 c.C991T
APC c.C1234T
KRAS c.G39C
SMAD4 c.C1421G
EGFR c.G2011A
Days after surgery
CC16035
ctDNA VAF (%)
TP53 c.G725T
PIK3CA c.G263A
APC c.C4132T
Days after surgery

continues

Supplementary Figure S2 conitinued

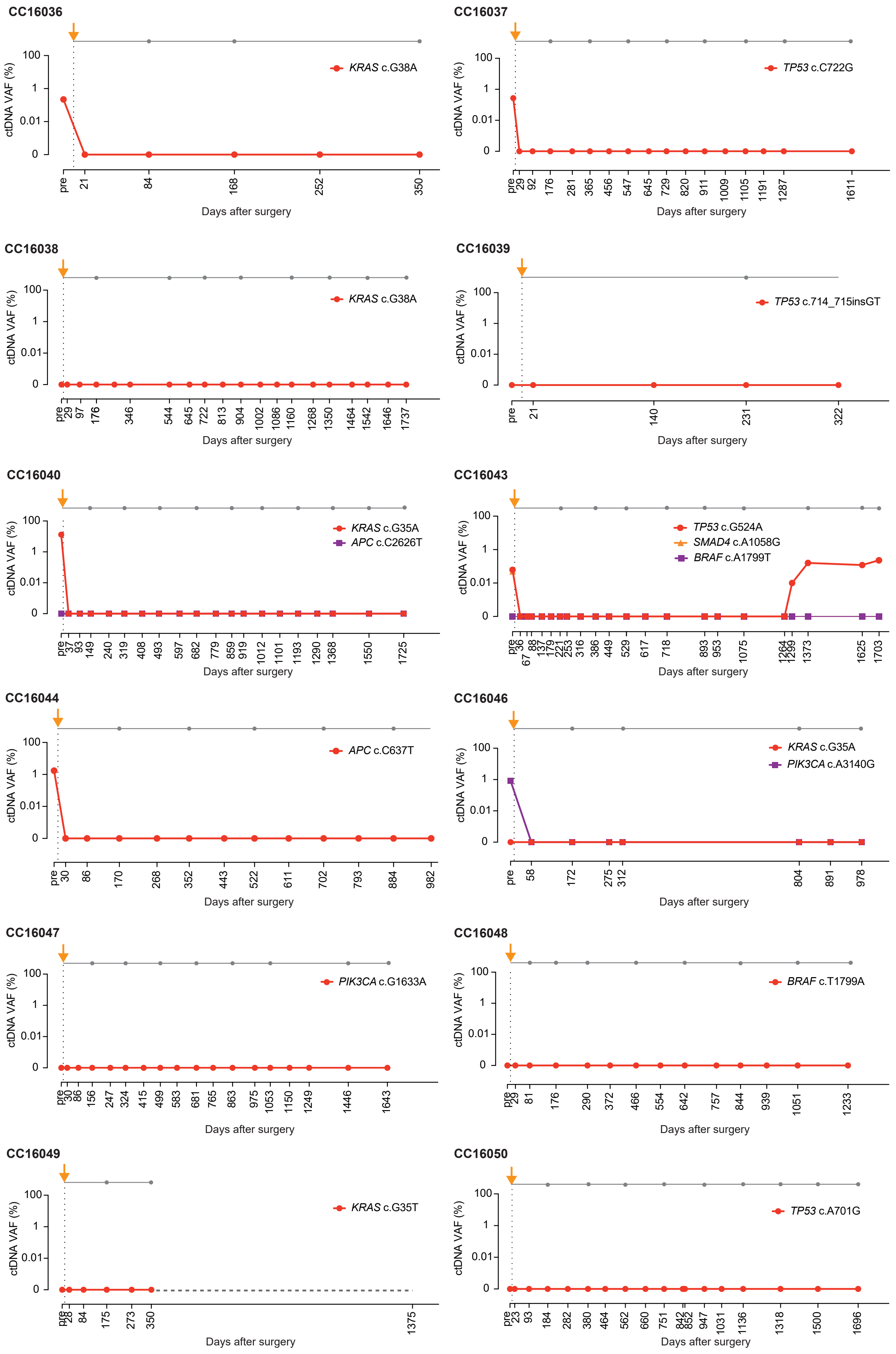


continues

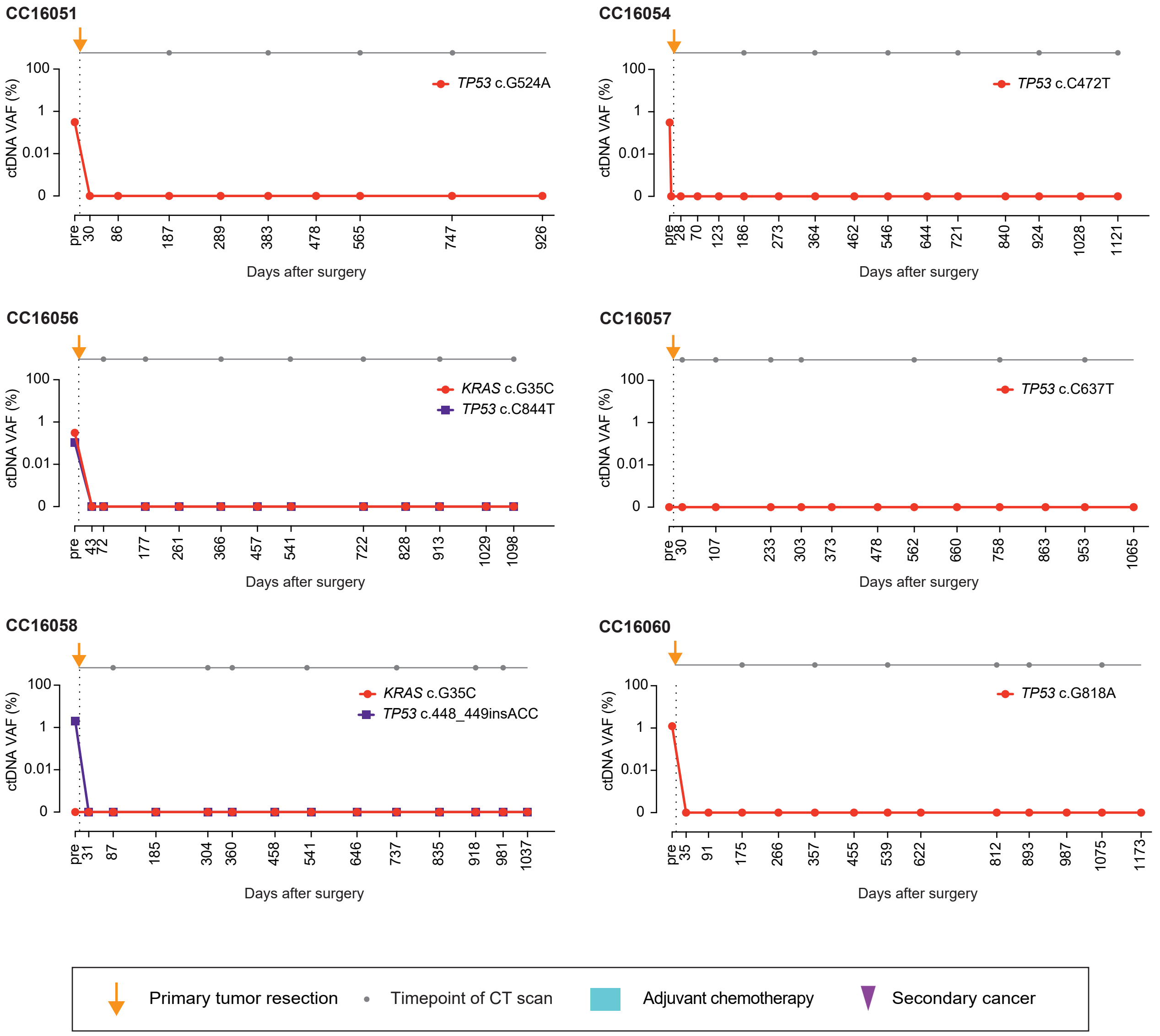


**Supplementary Figure S2. Dynamics of ctDNA in 42 patients with CRC without clinical relapse.** VAF, variant allele frequency.

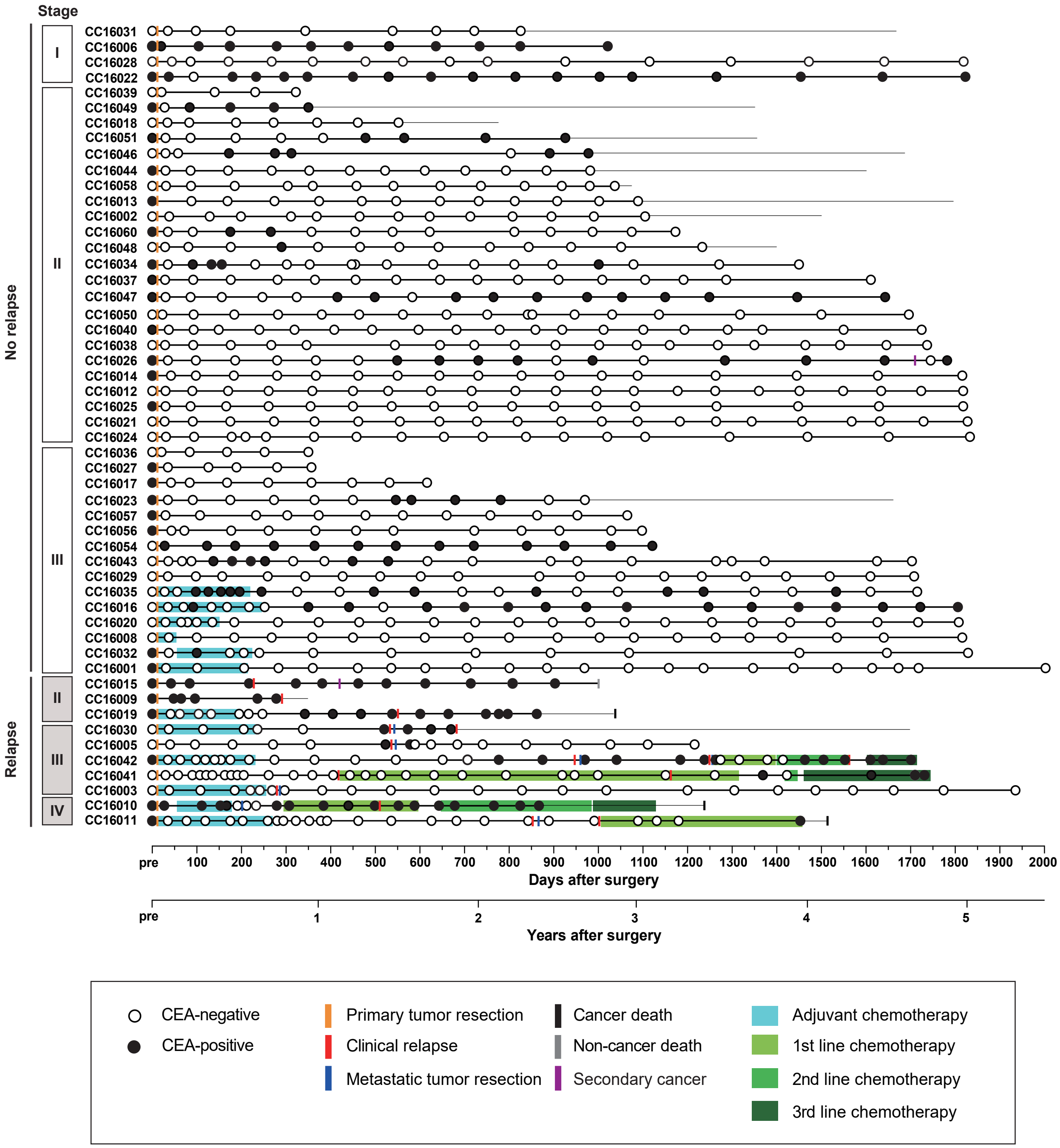


**Supplementary Figure S3. Longitudinal CEA monitoring of patients with CRC during the postoperative period.**
CEA, carcinoembryonic antigen.

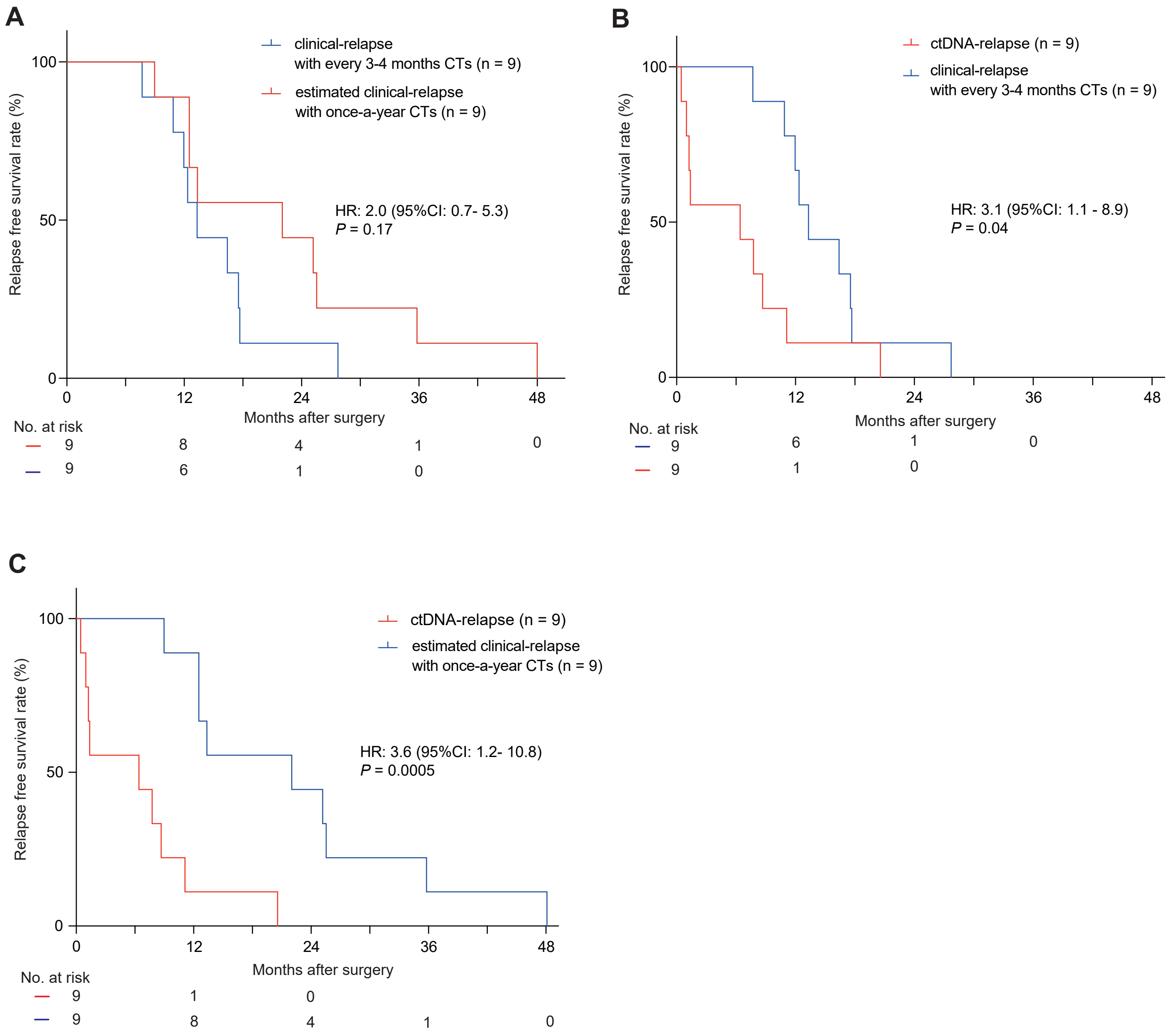


**Supplementary Figure S4. Comparison of relapse free survival (RFS) rates according to relapse type.**
(A) Clinical-RFS rates with CTs every 3-4 months and estimated clinical-RFS with once-a-year CTs. (B) Clinical-RFS and ctDNA-RFS rates. (C) Estimated clinical-RFS with once-a-year CTs and ctDNA-RFS rates. P values were derived using the Kaplan-Meier log-rank test. HR was calculated using the log-rank test. HR, ctDNA, circulating tumor DNA; CT, computed tomography scan; HR, hazard ratio.
